# An Agent-Based Simulation Using Extensive Real Datasets: the Case of COVID-19 in Catalonia

**DOI:** 10.1101/2024.07.10.24310130

**Authors:** M. Bosman, Y. Cordon, M. Duran, L. Gabbanelli, C. García-Pérez, X. Jordan, M. Manera, P. Masjuan, A. Medina, Ll.M. Mir, A. Oròs, V. Vitagliano

## Abstract

During the COVID-19 pandemic, effective public policy interventions have been crucial in combating virus transmission, sparking extensive debate on crisis management strategies and emphasizing the necessity for reliable models to inform governmental decisions, particularly at the local level. Leveraging disaggregated socio-demographic microdata, including social determinants, age-specific strata, and mobility patterns, we design a comprehensive network model of Catalonia’s population and, through numerical simulation, assess its response to the outbreak of COVID-19 over the two-year period 2020-21. Our findings underscore the critical importance of timely implementation of broad non-pharmaceutical measures and effective vaccination campaigns in curbing virus spread; in addition, the identification of high-risk groups and their corresponding maps of connections within the network paves the way for tailored and more impactful interventions.

## Introduction

The COVID-19 outbreak shed light on the critical role of timely and well-informed policy decisions in managing and mitigating the spread of infectious diseases. The challenge that policymakers have to face in an interconnected, global society is to find the correct balance between the economic and social impacts, together with psychological implications, of various interventions and public health considerations. Policies need to be adaptive, responding to quickly changing circumstances and emerging information. In this context, the significance of highly customizable simulations of epidemic models becomes evident: they provide not only valuable insights about the outbreak dynamics but also a versatile platform to promptly test scenarios in which different explicit containment measures (e.g., selective lockdowns, restrictions on specific mobility patterns, group-oriented vaccination campaigns) are put in place.

Mathematical modelling of sophisticated social environments can be consistently achieved within the framework of *agent-based models* (ABMs), computational models that simulate the emergent behaviour of complex networks starting from the structure of the interactions between the individual entities (the *agents*) of the system. Agents behave and interact with other agents and the environment in certain ways that would produce emerging effects that may differ from the effects of individuals. Concerning public health, this can be intuitively understood as the study of the spread of a certain disease – or, more in general, of unhealthy behaviours – in a community as a result of the demographic characteristics of single individuals and their social relations. The (abstract) control of the behaviour of each agent allows the evaluation of the response of the network to a given change and a relatively simple playground to identify the groups, or the links in the social network, where interventions could have the greatest impact^1^.

Epidemic ABMs can in principle provide a set of solution-focused tools to single out the most effective among various containment strategies. However, the robustness of the outcome of a simulation compared with the real spatiotemporal evolution of a disease is tightly entangled with the quality and volume of available socio-demographic data. To be realistic, the ABM-based simulation should rely on a network whose characteristics and properties reproduce, as closely as possible, the actual population. This implies having access to up-to-date granular repositories with high-resolution individual data, which, unfortunately, are not always available, rarely ready-made, and seldom public. Socio-demographic data provides a snapshot of the substratum in which the disease may propagate. The disease itself with its bio-medical characteristics plays as well a key role. This information must be incorporated in any attempt to spread modelling.

In this paper, we use *real, disaggregated census and mobility data* of the population of Catalonia with its *∼*8M people to build a network model and simulate the sequence of events that characterised the natural history of COVID-19 in Catalonia. To the best of our knowledge, our census dataset (including over 120 socio-demographic variables for a representative sample of 600k single agents in Catalonia) is one of the largest, raw datasets ever used to this scope.

The COVID-19 pandemic unfolded across the globe in early 2020, creating widespread disruptions in our societies. Many countries faced recurring waves of the virus, particularly intense in the initial two to three years. Stringent initial lockdown measures were followed by extensive vaccination campaigns that, together with the evolution of the virus into less deathly variants helped in restoring the situation to a level manageable by national health systems. Nevertheless, even four years later, new strains of the virus continue to circulate, posing ongoing challenges. Our time scope, which includes years 2020 and 2021, did not require us to consider evolving viruses and multi-strain overlapping waves, but ABMs allow relatively easy implementation of such an effect when necessary.

In a first paper^2^, we focused on the province of Barcelona, employing an ABM to track the contagion that originated from a small set of randomly chosen infected individuals in early 2020. Residence location, household structure, employment situation, and mobility routines, along with the resulting pattern of contacts, including incidental contacts such as those arising in public transportation or due to increased social activities during holidays, were inferred from detailed, disaggregated census data and information supplied by mobile network operators. The evolution of the disease in the host and its intensity were taken to be age-dependent and modelled according to the first observations available at the time. In the first phase of our work, we successfully reproduced the curve of diagnosed cases in 2020, highlighting the distinct characteristics of the two main waves based on individuals’ age and place of residence.

In the current simulation, *covering both 2020 and 2021 across all four provinces of Catalonia*, several improvements and additional features have been introduced. Notable enhancements include accounting for the impact of vaccination campaigns with different vaccines and a strongly age-dependent vaccination timeline. The wave patterns as revealed by epidemiological data varied among provinces due to differences in mobility, contacts, and the presence of distinct population groups affecting the disease propagation. Health personnel, residents in long-term care facilities, and workers in nursing homes have been treated separately given their roles during the outbreak. With these refinements, we have been able to simulate the five waves that occurred in 2020-21; we underscored the pivotal roles of lockdown measures and the vaccination campaign in controlling the pandemic and delved into the potential impact of different vaccine characteristics and vaccination timelines.

## Methods

The Basic Health Area (known as Àrea Bàsica de Salut, or ABS, in Catalan) serves as the fundamental territorial unit used by the Catalan Health Department for the organization of primary healthcare services in Catalonia^3^. Typically, each ABS caters to approximately 20,000 individuals and is linked to its respective network of hospitals and health proximity centres. Catalonia is home to 374 such areas, distributed across its four provinces, as depicted in Figure 1. The demarcation of these areas is influenced by a combination of factors, including geography, demographics, and social dynamics. Notably, close to major cities, especially Barcelona, ABS areas tend to be more compact with a higher population density.

**Figure 1.**
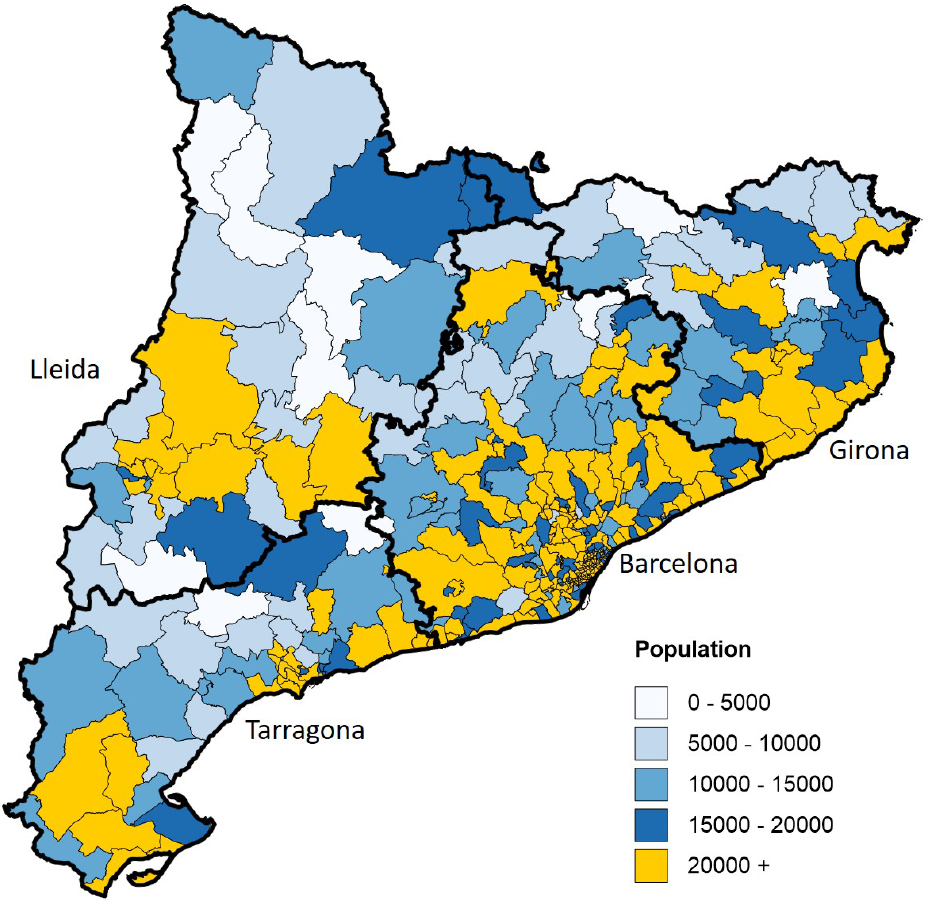
Map of Catalan ABSs. The map displays the 374 ABSs across the Catalan provinces of Barcelona, Girona, Lleida, and Tarragona (thick black lines); each ABS is depicted by a color code corresponding to its population size.

By integrating census data aggregated at the ABS level and daily mobility information between ABSs, we constructed a comprehensive model capturing realistic patterns of contacts and movements related to both work/school and social activities for the entire population. Healthcare system data on the daily counts of COVID-19 cases are used to calibrate our simulation. Information on the extensive vaccination campaign against COVID-19 launched in 2021 is also included.

### Census data

The “Cens de Població”, or population census, which provides socio-demographic information by categorizing the Catalan territory into 5,107 census sections, was made available upon request by the Spanish National Statistics Institute (Instituto Nacional de Estadística, INE^4^). The latest available census (2011) contains detailed information on around 10% of the population, covering aspects such as housing, education, work, family structure, etc. Considering the effective weight of each of these individuals, the dataset yields insights into the 7,472,937 inhabitants of Catalonia at that time. The original 2011 set has been reorganized to match the ABS structure (see Section 1 of the Supplementary Material (SM) for further information).

The census used for population reconstruction lacks information on individuals aged 65 years and above residing in nursing facilities. To address this gap, we consulted the list of 1,002 official long-term care facilities established in 2019, incorporating details on available spaces therein^5^. The age distribution among the elderly residing in these facilities displays an almost symmetrically inverse pattern compared to those in family dwellings^6^. The oldest individuals among the elderly predominantly reside in residential care facilities, while the younger ones live in private homes. An additional segment was introduced into the census file to accurately represent this specific demographic. Considering the average occupancy rate of nursing facilities at 86%^7^ and their respective locations, we reconstructed the demographic profile for an estimated population of 53,000 individuals residing in these homes. Ages were randomly assigned to individuals based on the overall age structure.

Additionally, during the summer season, Catalonia experiences an influx of temporary workers in the agricultural sector, with the province of Lleida hosting the largest proportion (see Section 1.2 in SM). We created an additional segment of the census file with over 4,000 temporary workers randomly assigned to mock farming companies. They reside in the same ABS as their workplace, share housing and are assigned social contacts like the rest of the population.

We devote special attention to two specific categories of sanitary workers (see Section 2.2 in SM). Sanitary workers engaged in geriatrics are estimated to be around 34,000, and sanitary workers operating in hospitals in close contact with infected patients at approximately 18,000.

### Healthcare system data

We obtained comprehensive and anonymized data on the daily counts of COVID-19 cases, hospitalizations, intensive care unit (ICU) admissions, and deaths through the Program of Data Analysis for Research and Innovation in Health (“Programa d’Analítica de Dades per a la Recerca i la Innovació en Salut”, PADRIS^8^). PADRIS operates under the auspices of the Agency for Health Quality and Assessment of Catalonia (“Agència de Qualitat i Avaluació Sanitàries de Catalunya”, AQuAS^9^). The data consist of two sets covering the period 2020-21: one providing the clinical history of individuals testing positive at least once (taken as a reference), and another with aggregated data by ABS and five-year age intervals. The latter includes details on the number of positive and negative test results, as well as information about the vaccination campaign categorized by age interval, along with specific details for nursing homes and healthcare workers.

Figure 2 illustrates the daily record of COVID-19 cases detected through PCR tests for the reference set. The data exhibit weekly fluctuations in the number of registered cases. These dips primarily result from reduced healthcare staffing and patients’ reluctance to seek medical attention for mild symptoms during weekends, leading to lower daily case counts across Catalonia. A noteworthy aspect is a disparity of approximately 10% between the two data sets, arising from their collection from different databases and variations in anonymization criteria. We recognize this as a systematic uncertainty in our analysis.

**Figure 2.**
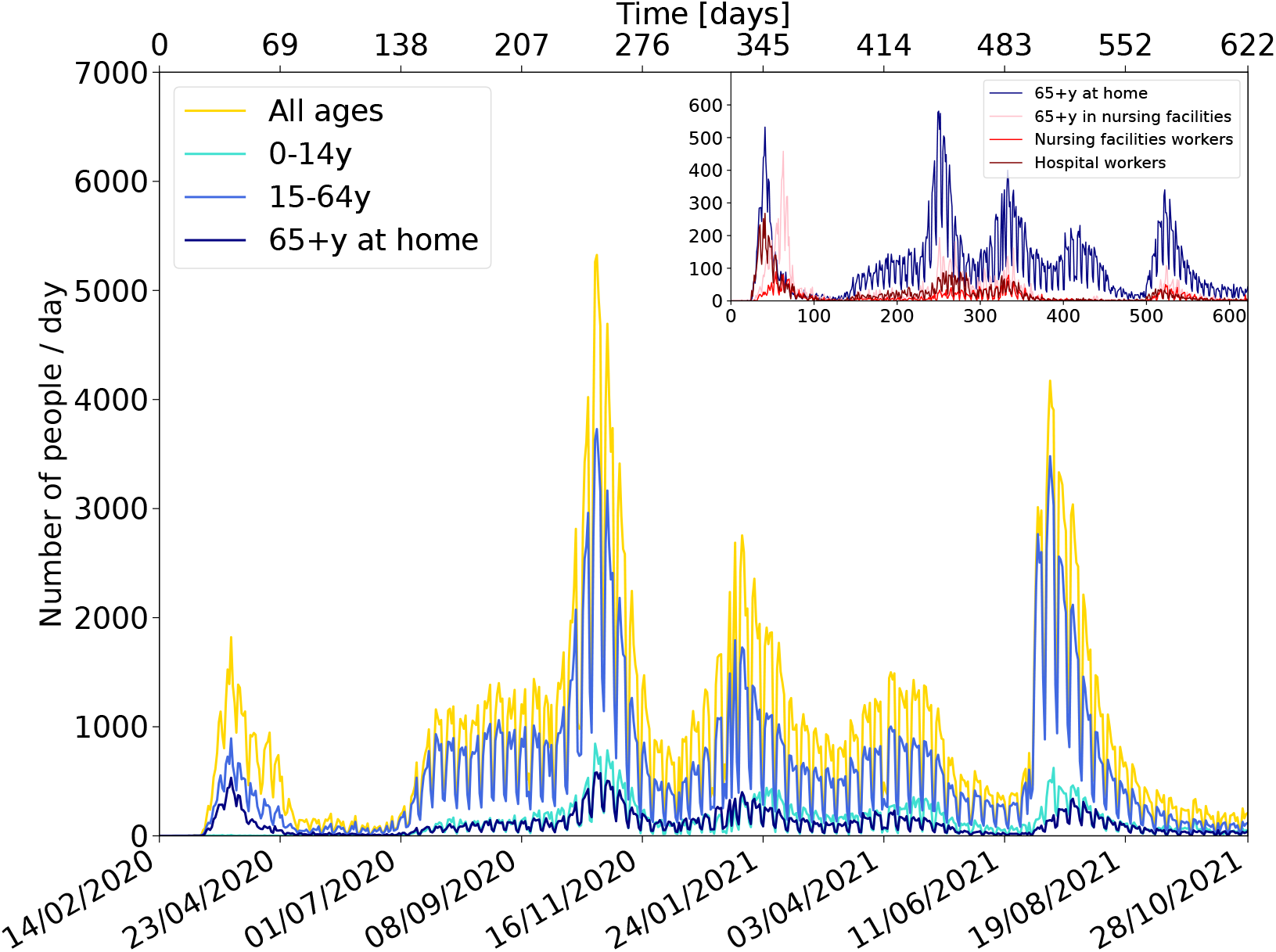
COVID-19 data in Catalonia. Total number of diagnosed people with PCR tests in Catalonia split by age categories: less than 15 years old, from 15 to 64 years old, and older (65+y) living at home. In the inset, people older than 64 years old living either at home or in nursing facilities, as well as nursing facilities and hospital workers, are shown.

### Mobility

In this study, we leverage two sets of processed mobility data sourced from the INE and from the Barcelona Supercomputing Center (BSC)^10^. Both datasets are derived from the analysis of the same raw data, detailing the positions of 80% of mobile phones with Spanish numbers over time, offering insights into population movements. Both studies quantify mobility based on trips between origins and destinations, with a minimum duration of 2 hours for INE and 20 minutes for BSC. INE attributes weekday mobility to work activities and weekend mobility to leisure activities. In contrast, BSC captures both work and leisure trips during both weekdays and weekends. BSC employs a general approach to project data across different geographical layers, going from higher granularity (*mobility areas* ranging from districts to municipalities depending on the density of population) to lower granularity (such as, in order, municipalities, ABSs, provinces, etc.). The highest precision is achieved by weighting the information based on the number of inhabitants, available in the form of a 1 km^2^ grid from GEOSTAT^11^. In this work we use the mobility data from the BSC projected on the ABSs and those from the INE averaged for all of Catalonia.

Figure 3 illustrates the weekly evolution relative to a pre-COVID-19 reference week for both sets of data aggregated over Catalonia. The mobility variation pattern is correlated between the two datasets and shows a consistent alignment with lockdown measures and holiday periods, as previously explored in our study^2^ and discussed by BSC^12^. While the level of mobility is similar for work/school activities, there are notable differences in leisure activities. BSC conducted a comparative analysis of their data with that of INE^10^. On average, BSC reports approximately ten times more trips than INE: the difference has to be traced back to the two distinct definitions of trips previously detailed, requiring longer stays in the case of INE. The correlation remains robust, with a Pearson’s coefficient close to one when aggregating over larger areas like Catalonia, but slightly diminishes to about 0.8 when comparing data from smaller geographic areas.

**Figure 3.**
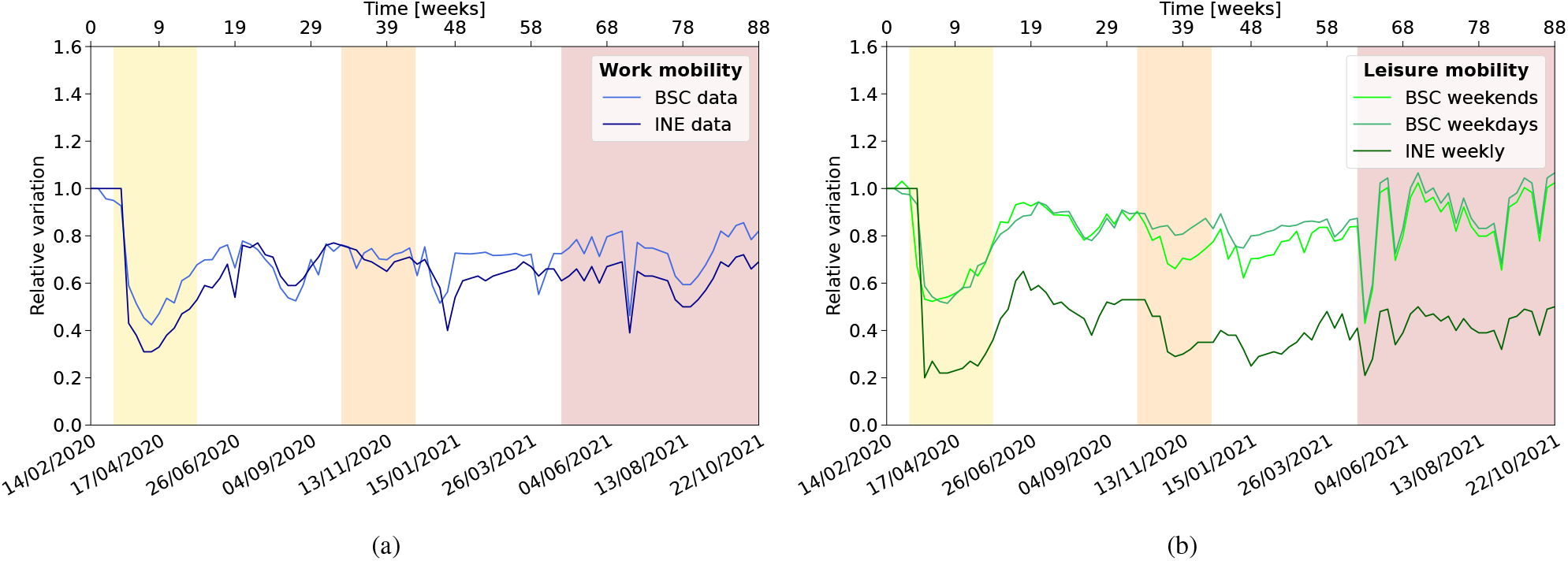
Mobility Evolution. The average daily mobility in Catalonia for **(a)** work/school activities and **(b)** leisure activities during weekdays and weekends, from INE and BSC analyses. Shaded vertical bands highlight periods of varying mobility restrictions, corresponding to the first, second, and fourth/fifth waves of the pandemic.

The ratio between the average daily mobility derived from BSC and INE datasets is close to one for work activities but increases to around two for leisure activities. Moreover, in correspondence with the outbreak peaks (periods characterized by stricter lockdown measures), the ratio tends to be higher, indicating a more pronounced reduction in longer trips compared to shorter ones (see SM Figure S.2). Our model incorporates mobility information in two fundamental ways: firstly, to approximate the impact of containment measures on people’s contact patterns, and secondly, to delineate various population characteristics, as elucidated in the next sections. We hypothesize that a decrease in mobility corresponds to a reduction in the viral load to which individuals are exposed, although the precise effectiveness of this reduction remains uncertain. To address this uncertainty, we have introduced a calibration factor matched to data that translates the level of mobility into an estimate of the reduction in effective viral load, which in the case of leisure activities depends on the level of restrictions.

### Workplaces, schools, and places for leisure activities

Each member of the population is assigned a workplace or a school/university, if applicable, as well as a location for leisure activities based on their age and information obtained from the census. Census data provide insights into the occupational category of individuals, which we categorize into six sectors: primary sector, industry, construction, services, education, and healthcare. IDESCAT^13^, drawing from data in the “Directori central d’empreses” (DIRCE)^14^, furnishes details about the size and distribution of companies per sector. Schools typically consist of 30 classes of varying sizes, depending on age (0-18y) (see ref.^15^ and table S.11). Synthetic schools are established per ABS to accommodate the corresponding number of pupils living therein. University campuses are established based on official data regarding the location and the registered number of students^16^. Similarly, nursing homes and hospitals are created according to their respective locations and bed capacities^5,17^.

We use mobility data to discern patterns of movement between different ABSs for work/school and leisure activities. For trips from home to work, we identify, for each ABS, the corresponding list of target work ABSs ranked by frequency and distance. Census data provide information on the duration it takes for individuals to commute to work (or for children to travel to school), as well as their mode of transport. This is translated into distance, and a destination ABS could be in principle chosen accordingly. In the case of work/school, we distinguish two cases: companies for which the exact ABS and size are known (e.g. universities or residencies) and those for which these data are not known. In the first case, we assign the company to workers according to distance; otherwise, we use census data to simulate a geographical distribution of businesses and educational institutions, and accommodate the list of workers of the corresponding ABS. ABSs visited for leisure activities during weekdays and weekends are allocated to single individuals based on the ABSs list provided by mobility data.

The BSC data enables monitoring of the total population residing in ABSs over time. Significant population movements are observed during the summer, with approximately 0.4 million people departing from Barcelona to visit Mediterranean coast resorts and other destinations. This is factored into the simulation, resulting in adjustments to the set of leisure contacts accordingly.

### Vaccines and vaccination campaign

In 2021, Spain launched an extensive vaccination campaign against COVID-19. The campaign commenced in January, prioritizing healthcare workers, followed by subsequent rollouts organized by age groups from oldest to youngest^8,13^. Participation in the vaccination drive was voluntary, and the level of uptake was notably high, exceeding 90% for individuals over 45 years of age, albeit slightly lower among younger demographics. Children under 12 were not eligible for vaccination.

The campaign administered four different vaccines belonging to two types: mRNA-based vaccines including Pfizer-BioNTech^18^ and Moderna^19^, and viral-based vaccines such as AstraZeneca^20^ and Janssen^21^. While the majority of individuals received mRNA-based vaccines, those in the 60-69 age group were primarily vaccinated with viral-based vaccines. The vaccination process typically involved administering a first dose followed by a second dose one month later (or two months for viral-based vaccines), followed by a booster dose six months later. Figure 4 illustrates the vaccination profile, depicting the distribution of first, second, and eventual third doses across the entire population, as well as aggregated within various age categories, according to PADRIS data.

**Figure 4.**
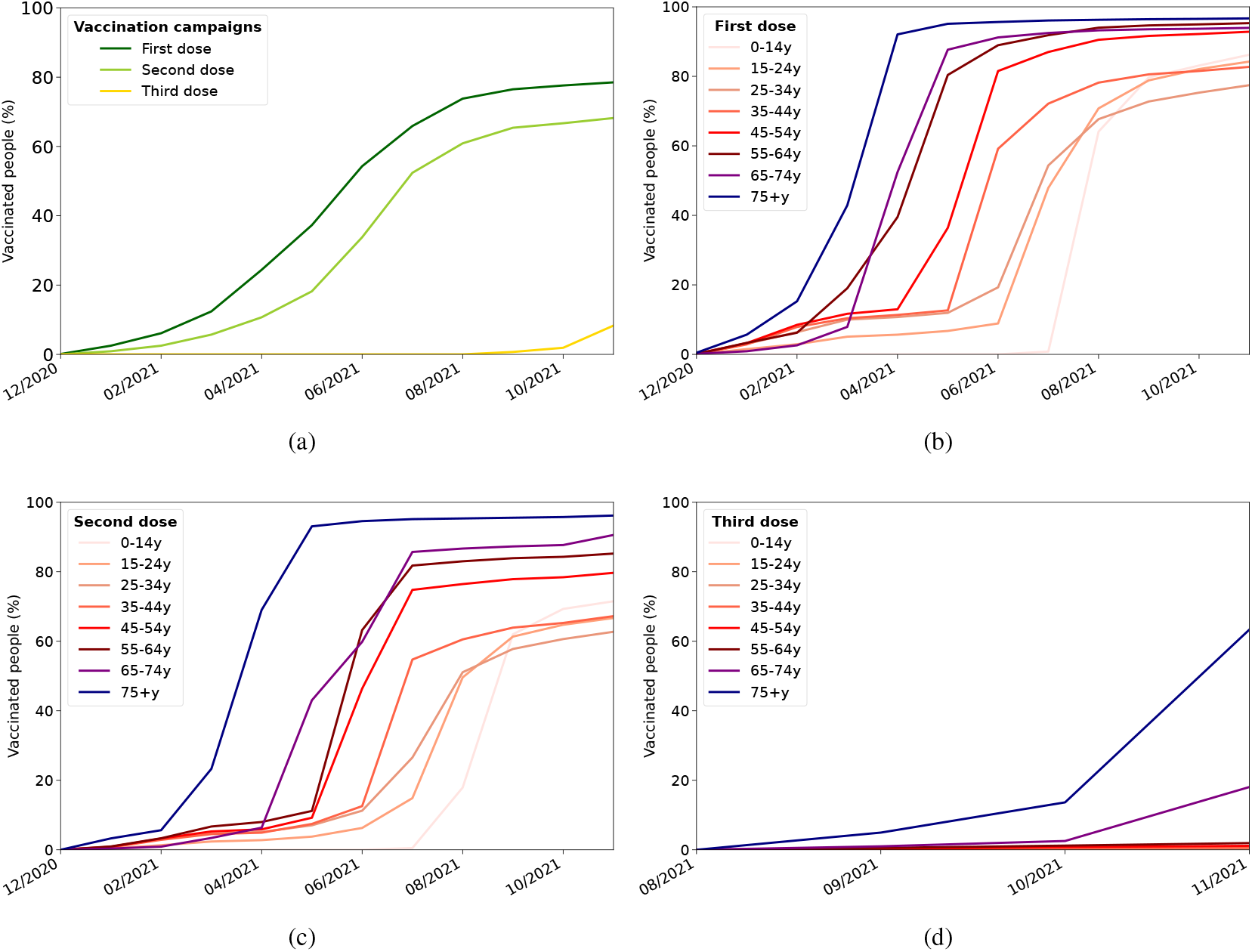
2021 vaccination campaign in Catalonia. **(a)** Time profile of vaccination campaign in 2021: the first dose started to be administered in January, the second dose one month later and the third dose six months later. **(b)** Administration of the first dose, **(c)** second dose, **(d)** third dose, by age category.

The effectiveness of the vaccines is inferred from published data^22^. Figure 5 delineates the two effects of the vaccine considered in the simulation: the reduction of the probability of infection and the attenuation of symptoms with a corresponding decrease in viral load emission. For mRNA-based vaccines, the efficacy (contagion probability reduction) stands at 47% after one dose and rises to 92% after two doses. This effectiveness remains stable for four months before gradually declining to 47% during three months. On the other hand, viral-based vaccines exhibit 40% efficacy after one dose, increasing to 76% after two doses. This efficacy remains steady for three months before decreasing to 40% during three months. All booster doses administered are of the mRNA type, reinstating efficacy to 92%, which remains constant throughout the simulation period. Additionally, the reduction in viral load shedding, associated with symptom alleviation and disease severity, results in a 10% reduction for every administered dose^23^.

**Figure 5.**
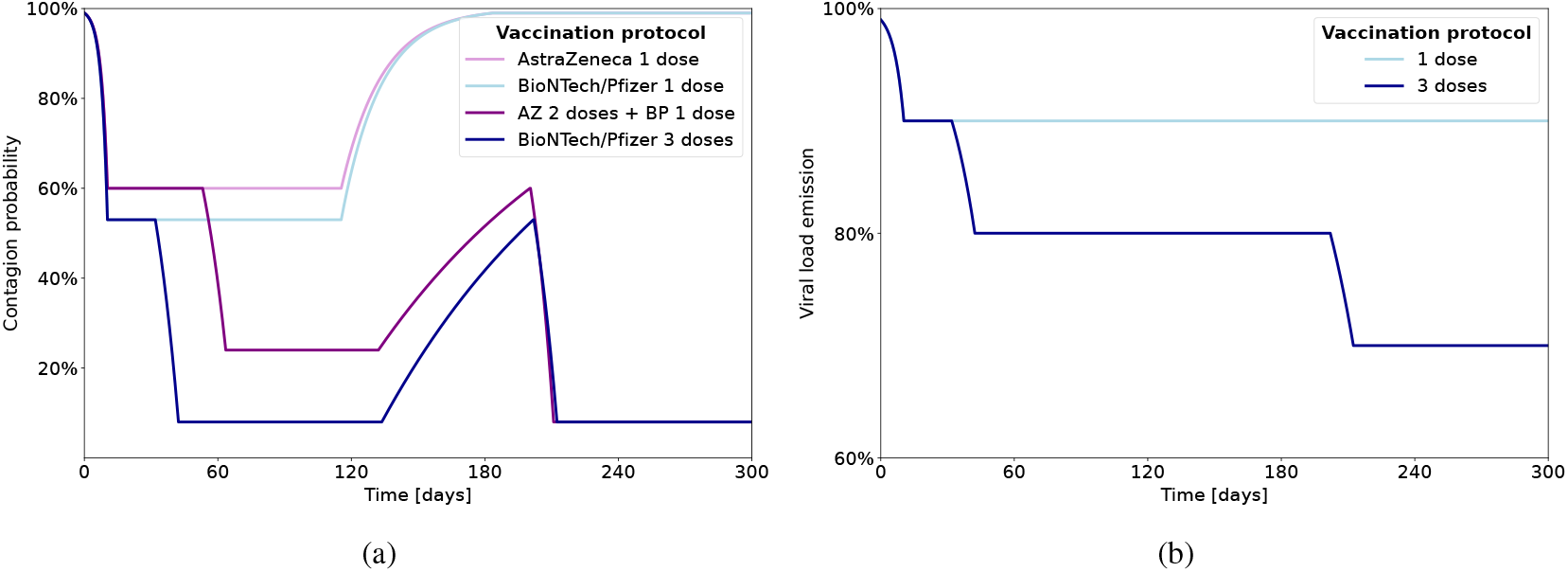
Effects of vaccine. The vaccine has two effects: **(a)** Reduction of contagion probability due to vaccine. The reduction of probability of becoming infected is shown for two cases: a single dose of vaccine, and three doses of vaccines for Pfizer/Moderna and AstraZeneca/Janssens. **(b)** Moderation of the infectious process with the corresponding reduction of viral load emission.

Tables S.8 and S.13 in the SM provide comprehensive insights into the model for the vaccination campaign – encompassing age categories, vaccine types, initiation dates, intervals between doses, and population coverage for each dose –, as implemented by default in the simulation. Each age category required approximately 40 days for complete vaccination. Since details regarding the administration of the third booster dose are lacking, we presume that individuals who received two doses eventually received a third. This assumption underpins the simulation’s continuity and ensures a comprehensive representation of vaccination dynamics. We have tested several scenarios, exploring diverse vaccine efficacies and timelines of administration to assess their impact on outbreak evolution.

### Model design for the COVID-19 spread

In our model, each individual in the population of Catalonia is assigned to one (and only one) of the following compartments at any given time: *susceptible, exposed, infected, diagnosed, dead, recovered*, and *immune* (note that in our model we do not include traditional births dynamics). When susceptible individuals come into contact with infected persons, their state may transition to exposed based on the probability

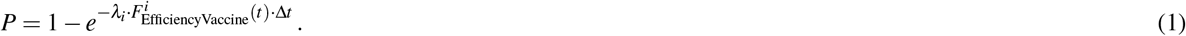

Here, the force of infection, *λ*_*i*_, represents the total viral load a *single individual i* is exposed to per unit time (day) 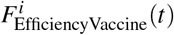 denotes the reduction in the risk of infection resulting from vaccination, and Δ*t* is the time interval (1/3 day). A comprehensive mathematical description of the estimate of *λ*_*i*_ is given in Section 2.1 of the SM. Here we will limit the discussion to a general description of the computational strategy.

The total viral load exposure *λ*_*i*_ is a composite of exposures occurring throughout the day across various settings. It primarily encompasses contributions from three distinct eight-hour intervals corresponding to an individual’s time spent at home, work, or school, and engaging in social activities. To compute *λ*_*i*_, we calculate the viral load emitted by every individual and multiply by matrices describing the network of contacts. Additional contributions are considered for individuals using public transportation or visiting particularly crowded areas during their daily routines (for example, tourist areas during summer, or commercial areas during season holidays). In that case, to compute *λ*_*i*_, we estimate the average viral shedding of people involved in these activities or encountered within these settings, modulated by ABS-dependent mobility. We then multiply by the number of estimated contacts, e.g. during a typical trip in public transport, or the number of additional contacts during leisure activities (the latter are ABS-dependent and typically higher in summer coastal resorts or tourist areas of Barcelona). Details about relative contributions are given in Section 5 of the SM.

After the exposure, our population model considers personalized disease progression for each individual, with characteristics such as age-dependent symptoms and viral shedding levels (a survey of the epidemiological data used in the model is available in a previous publication^2^). These factors influence other outcomes, such as the likelihood of diagnosis or hospitalization. Infected individuals are classified based on symptoms (symptomatic or asymptomatic) and viral shedding intensity (strongly infectious, moderately infectious, or non-infectious). Three key combinations emerge: asymptomatic non-infectious (ANI), asymptomatic moderately infectious (AMI), and symptomatic strongly infectious (SSI). Age plays a crucial role, with older individuals more likely to exhibit symptoms and higher infectiousness, while children tend to be ANI. Additionally, symptomatic individuals are typically twice as infectious as asymptomatic ones. The overall infectiousness level is set for every individual according to these categories.

The model also incorporates probabilities of hospitalization and intensive care unit (ICU) admission, which correlate with symptom intensity. However, detailed temporal dynamics post-diagnosis, including hospitalization progression, are not explicitly modelled. Upon diagnosis, a portion of the population is tagged as hospitalized or in ICUs. Death can occur regardless of diagnosis or hospitalization, while recovery follows a fixed time frame unless death intervenes. Recovered individuals are considered immune against further infection for an average of nine months, with a root mean square (RMS) deviation of three months. In the case of a reinfection, their viral load emission is reduced (see Table S.6).

### Model Calibration

Our simulation model is constructed upon 194 parameters to account for the population description, the disease characteristics, the modelling of contacts, and the vaccination campaign (see Section 6 of the SM for a compilation). Most of the parameters can be set *a priori* from existing data (see Subsection 3.1 in the SM). Nevertheless, given that some of them are poorly known and difficult to determine precisely only from this comparison, we study the individual sensitivity of the model to each parameter and further calibrate the model via a goodness-of-fit test algorithm.

Ideally, to reproduce more accurately the observed evolution of diagnosed people, a simultaneous fit of all parameters should be performed (cf. Subsection 3.3 of the SM). However, this would require a complete modelling of the correlations between all parameters, with their respective ranges of variation, for which there is not enough knowledge. We follow instead an approximate procedure, concentrating on the most sensitive parameters. The fits are done successively one at a time with their respective systematic and statistical uncertainties. The cost function for each parameter is based on a *χ*^2^ statistic.

Only the three most sensitive parameters – broadly affecting age, spatial and time dependence – are calibrated using this procedure; in decreasing order of sensitivity, these are related to the mobility of people for leisure activities, the global infectiousness of the virus, and the relative weight of this infectiousness across different age groups. The calibrations are performed over the first year of the evolution of the disease, before the data and model are directly influenced by the active vaccination campaign. Additionally, due to shortcomings in the real-life data collection process, it is not possible to perform comparisons on a daily basis; instead, data has to be aggregated on a weekly basis, taken from Friday to Thursday to account for delayed registers. Further details on these considerations and the calibration process can be found in Section 3 of the SM. We estimate a minimum of 10% relative uncertainty to be associated with the calibration of these three parameters, which is represented in the figures by a shaded area. This does not include the uncertainty that may originate from imperfect knowledge of the other parameters.

## Results

### The natural history of COVID-19 in Catalonia

Figure 6 shows the results of our simulation after the model calibration process, extended to the full two-year evolution and compared against the collected data, aggregated across all provinces and age groups. The results span from February 14, 2020, to the end of October 2021, the period with consistent data availability.

**Figure 6.**
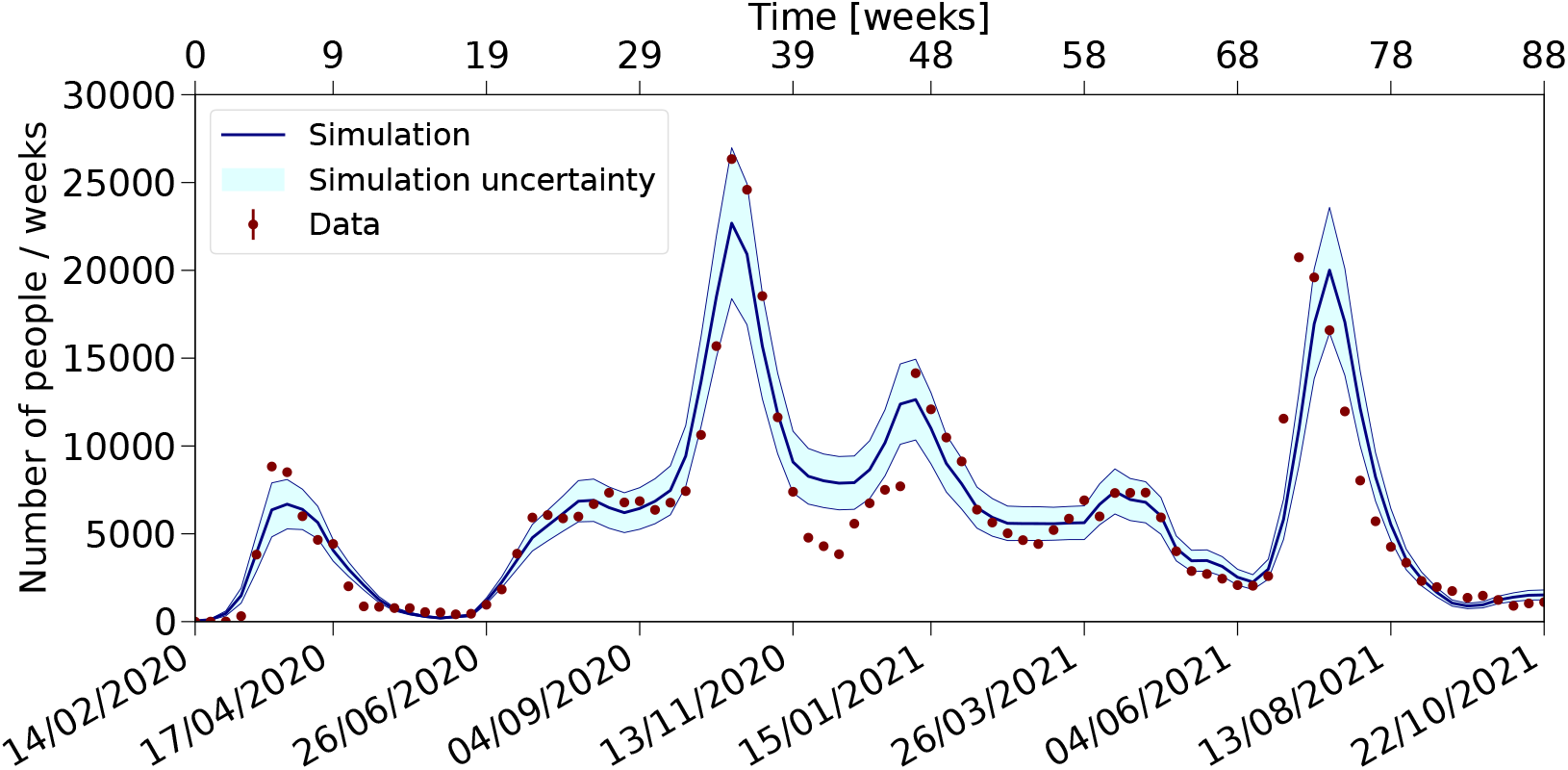
Number of diagnosed people in Catalonia. Data on diagnosed people are compared to simulation results for the period from 2020 to 2021.

During 2020, Catalonia experienced two distinct waves. An initial wave in March, whose shape is influenced by the specific characteristics of the disease and the mobility trends. Among these factors, the force of infection, pre-symptomatic viral shedding, and disease duration stand out, as well as changes in mobility patterns and the gradual recovery of work-related mobility. Thanks to the strict lockdown measures implemented, this initial outbreak was mitigated, and it was followed by a plateau. The summer plateau’s level and shape are linked to post-lockdown contacts, especially summer activities, and their timing. As lockdown restrictions eased and activities resumed — partially at first, then almost fully — after the summer, the increase in disease incidence at the end of this season prompted the emergence of a second wave in October. This latter wave was effectively curtailed by ad-hoc lockdown measures.

In 2021, the vaccination campaign played a crucial role in managing subsequent waves, although three additional waves were observed, each triggered by social gatherings during holiday periods: Christmas, Easter break, and summer holidays. These waves, including those in January 2021, are similarly shaped by changes in contacts, with the impacts of the vaccination campaign becoming increasingly apparent.

The analysis of the different components of the viral load *λ*_*i*_ (see SM Fig S.15) shows that the contribution from “home” dominates, especially in the periods of strong confinement. In the periods of more mobility, the “leisure” contacts are the second most relevant, followed by “work” activities.

Similar results for more specific subgroups (children, adults, seniors, nursing home residents and workers, and hospital workers) are presented in the SM (see Figs. S.5-S.10), highlighting differences in symptomatology and testing patterns across waves. Children are mostly asymptomatic, while old people are mostly strongly infectious. During the first wave only people with strong symptoms were tested, so very few children were diagnosed. Later on, a broader spectrum of people were tested, including close contacts of diagnosed people.

We also compare our model against data aggregated by provinces (SM, Figs. S.11-S.14). The Barcelona province, hosting the largest fraction of the population and with the strongest statistical power in the fit, is well reproduced in the simulation, along with the main features of the other less populated provinces. All three provinces exhibit essentially the same five waves as Barcelona, albeit with some differences in relative intensity. Notably, the summer 2020 “plateau” observed in Barcelona is absent in Girona and Tarragona, where a more gradual increase is observed. Additionally, Lleida featured an additional strong wave in July 2020 associated with the influx of temporary workers in the agricultural sector. The correlation between the daily evolution of waves in different provinces provides insights into their nature, as previously discussed^24^. Pearson’s correlation coefficients between diagnosed cases in the four provinces during the March and October waves exhibit a high degree of correlation, reflecting the synchronous spread of the virus (SM, Table S.4). However, during the summer period, characterized by increased holiday activities and foreign visitors, correlations are weaker, and in the case of Lleida, even negative due to specific local factors such as the influx of temporary workers in agriculture. The simulation generally reproduces the observed correlation patterns, indicating its capability to capture the essential features of disease spread within the Catalan territory.

Estimates of contacts across provincial borders during leisure activities reveal higher exchange rates between Barcelona and its neighbours (see Table S.5 of the SM). This exchange disproportionately affects less populated provinces, with significant impacts during summer as residents from Barcelona (about 400k people) travel to Mediterranean coastal resorts and the Pyrenees region. The relative impact in the provinces of Girona, Tarragona, and Lleida is 30, 20, and 10%, respectively. This effect is incorporated into the simulation, virtually reallocating part of the population in different ABSs during summer, which also implies changing the list of potential contacts. In any case, the tendency for the simulation to overestimate disease incidence in the outer provinces is possibly due to assumptions about contact patterns not fully accounting for differences in population density.

### The 2021 vaccination campaign

The prompt start of the vaccination campaign in 2021, along with its age-dependent profile and high level of participation, played pivotal roles in controlling the virus’s spread, limiting the number of infected cases, and facilitating the relaxation of containment measures to revive economic activities (details of the modelling of the vaccination campaign are collected in Table S.13 of the SM). Figure 7 demonstrates the significant impact of the vaccine campaign: in particular, the number of diagnosed cases, which would have shown a high peak during the summer if no vaccination measures were implemented, was instead reduced to a manageable level even while mobility was at its highest. The timeliness of the campaign was crucial, as a delay of three months would not have entirely prevented the summer peak but would only have decreased its severity. Such a scenario would likely have required the enforcement of stringent lockdown policies, with an adverse impact on society. Thus, vaccination emerged as a crucial component in the journey back to “normality”. We explored a scenario where vaccine effectiveness was reduced and assumed all vaccines administered were the same, with a 76% reduction in the probability of infection. This situation led to three times more diagnosed cases, underscoring the importance of vaccine efficacy in controlling disease transmission.

**Figure 7.**
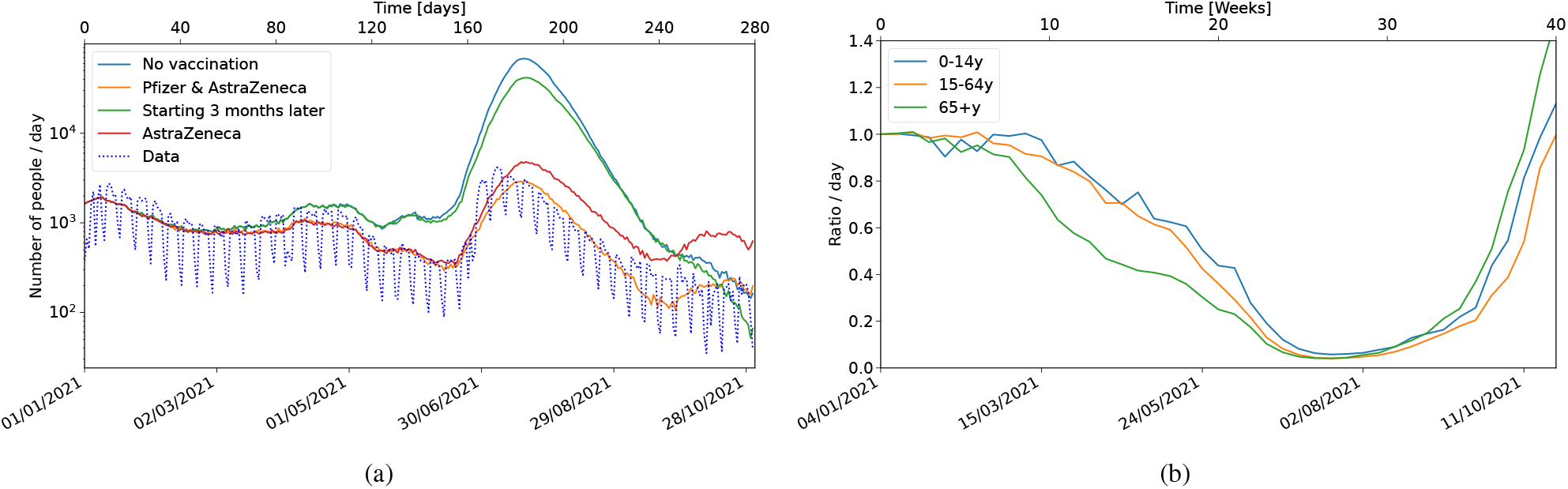
Diagnosed people for different vaccination scenarios. **a** Number of diagnosed people in 2021 in a no-vaccination scenario compared with different vaccination campaigns, including the actual campaign implemented in Catalonia. **b** Ratio of the number of diagnosed people in 2021 with and without vaccination for the vaccination campaign implemented in Catalonia for different age categories. The 15-64y category includes hospital and residency workers, and the 65+y includes residents in nursing homes.

### Data limitations

While the census data provide a detailed description of the population, including home composition, unavoidable simplifications arise due to data limitations. First, and more importantly, the reliability of the recorded number of positive cases is severely mined by under-reporting^25–27^ (for both infections and deaths). In the second instance, mobility data obtained from mobile phones lack age information and do not offer objective insights into age group differences. Furthermore, while mobile phone traffic provides geographic displacement data both for leisure and work-related activities, information regarding workplace size, location, and company type was unavailable at the desired granularity. Nonetheless, our analysis highlights the timely implementation of containment measures and vaccination campaigns by authorities as crucial factors in controlling epidemics.

## Conclusions

We have developed an advanced agent-based simulation model tailored to accurately reproduce the dynamics of COVID-19 spread in Catalonia throughout 2020 and 2021. This comprehensive simulation encompasses all the essential ingredients with enough precision to reproduce the flows of the pandemic across various age groups and provinces over the entire period under study. Our approach relies on high-quality disaggregated data, and not only provides valuable insights into spatial autocorrelation concerning the COVID-19 incidence during different phases of the outbreak but also estimates the impact of external interventions on human behaviour.

Several strengths of our method are worth highlighting. First and more importantly, our use of a granular representation of the population: the consistent availability of mobility, census, and health data at relatively small spatial units (ABSs) makes possible a robust calibration and comparison with real-world data, allowing for the discovery of important factors causing disease transmission; our agent-based model avoids the drawbacks of averaging population characteristics over broad regions and provides a more realistic description of local dynamics. Accurate modelling of contact patterns is ensured by the granularity of mobility data, which takes into consideration seasonal fluctuations and their impact on the virus spread. In this way, we are able to successfully capture both the effects of varied lockdown measures across different regions and the movements of populations with high temporal and spatial resolution.

The combination of health data with our model provides also a faithful replica of the age- and time-dependent vaccination campaign, which is a crucial aspect for understanding changes in the population behaviour that occur during the second year of the outbreak. The model can be easily expanded to include additional (epidemiological and etiological) virus characteristics, as well as demographic factors. Furthermore, although initially developed for Catalonia, our simulator can be adjusted for analysing other contexts at different geographical scales, upon the availability of high-quality data.

In conclusion, thanks to its accuracy, our model can serve as a reliable tool for assessing the efficacy of containment measures (in particular at a local scale) and providing invaluable insights to delineate targeted public health strategies.

## Data Availability

The population census, was provided by the Spanish National Statistics Institute (Instituto Nacional de Estadística, INE). The health data were provided by PADRIS (Programa d'Analítica de Dades per a la Recerca i la Innovació en Salut) operating under the auspices of AQuAS (Agència de Qualitat i Avaluació Sanitàries de Catalunya). In compliance with European and national laws, the above datasets were only made available to the researchers participating in this study and cannot be shared by them with other parties. Researchers can request census data from INE at https://www.ine.es/ss/Satellite?c=Page&p=1254735550786&pagename=ProductosYServicios%2FPYSLayout&cid=1254735550786&L=1, and from AQuAs by contacting PADRIS at. The sets of processed mobility data are publicly available from INE at https://www.ine.es/experimental/movilidad/experimental_em4.htm, and from BSC at https://github.com/bsc-flowmaps. Data sets generated during the current study are available from the corresponding author on reasonable request.

https://www.ine.es/experimental/movilidad/experimental_em4.htm

https://github.com/bsc-flowmaps

## Acknowledgements

The authors affiliated to CED, IFAE and i2CAT acknowledge the support of the CERCA institution, Centres de Recerca de Catalunya. They acknowledge the support of the “Agència de Gestió d’Ajuts Universitaris i de Recerca” (AGAUR) via the grant PANDE00180 “A powerful stochastic tool to assess the impact of the COVID-19 in Catalonia integrating detailed demographic and mobility data” of the program PANDÈMIES 2020 “Replegar-se per créixer: l’impacte de les pandèmies en un món sense fronteres visibles”. The grant funded the work of YC, AO and (partially) of AM. The authors acknowledge the support of PADRIS (“Programa d’Analítica de Dades per a la Recerca i la Innovació en Salut”) operating under the auspices of AQuAS (“Agència de Qualitat i Avaluació Sanitàries de Catalunya”) for providing the health data. They acknowledge the help of Albert Esteve from CED-CERCA in obtaining the PADRIS and Census data.

The work of VV has been partially funded by Next Generation EU through the project “GeTOnQuaM”. The research activities of CGP and VV have been carried out in the framework of the INFN Research Project QGSKY. VV extends his appreciation to the Italian National Group of Mathematical Physics (GNFM, INdAM) for its support.

## Author contributions statement

MB led the conception of the project. YC, MD, LG, CG, MM, LLM, PM, AO and VV contributed to the modelling design. MB, YC, AM, AO took care of data preparation. MB, YC, MD, LG, CG, MM, LLM, AO developed the code. MB, PM, CG, LLM, VV wrote the manuscript. All authors contributed to the discussion and interpretation of the results, revised critically the draft and approved the final version of the manuscript.

## Additional information

The authors declare no competing interests.

## Data availability

The population census, was provided by the Spanish National Statistics Institute (Instituto Nacional de Estadística, INE). The health data were provided by PADRIS (“Programa d’Analítica de Dades per a la Recerca i la Innovació en Salut”) operating under the auspices of AQuAS (“Agència de Qualitat i Avaluació Sanitàries de Catalunya”). In compliance with European and national laws, the above datasets were only made available to the researchers participating in this study and cannot be shared by them with other parties. Researchers can request census data from INE at https://www.ine.es/ProductsAndServices/StatsticalInformation.cat, and from AQuAs by contacting PADRIS at. The sets of processed mobility data are publicly available from INE at https://www.ine.es/experimental/movilidad/, and from BSC at https://github.com/bsc-flowmaps. Data sets generated during the current study are available from the corresponding author on reasonable request.

