## Supplementary Material for "An Agent-Based Simulation Using Extensive Real Datasets: the Case of COVID-19 in Catalonia"

The supplementary material includes additional information on the external input data, the model itself, the parameter settings and calibration procedures, the resulting predictions, the components of viral load, and tables with parameter values.

#### 1 External input data

##### 1.1 Census data

The "Cens de Població", or population census, provides socio-demographic information by categorizing the Catalan territory into 5,107 census sections. These data need be reorganized into the 374 Basic Health Area (known as Àrea Bàsica de Salut, or ABS, in Catalan)[3].

To achieve correspondence between areas with differing borders, we used a geo-processing technique, computing the centroid of each census section. Through this method, each census section is linked to a specific ABS. Although certain census sections may be associated with two or more ABSs, the centroid consistently corresponds to a single ABS. Given the small size of census sections, this approach ensures sufficient precision.

After establishing the correspondence between census sections and ABSs, we employed the raw statistical census data corresponding to the 374 ABSs, provided on request by the Spanish National Statistics Institute (Instituto Nacional de Estadística, INE[4]). Due to sampling limitations and the need to preserve anonymity, INE does not provide detailed data for regions with populations of less than 20,000 inhabitants. Since only 180 ABSs have populations exceeding this threshold, we needed a regrouping strategy for the remaining 194 ABSs. This regrouping was guided by three criteria: ensuring territorial continuity, combining ABSs from the same county, and preserving as many of the 180 larger ABSs as possible. This approach resulted in a total of 246 regions with populations exceeding 20,000 inhabitants.

The file from the latest available census of 2011 contains detailed information on over 120 variables of individual data, covering aspects such as housing, education, work, family structure, etc., for approximately 600,000 individuals (around 10% of the population). By considering the effective weight of each of these individuals, the dataset yields insights into the 7,472,937 inhabitants of Catalonia at that time. Table S.1 presents the statistics of various subsets of the population, as implemented in the simulation.

The census data were published in 2011, while our study focuses on the propagation of COVID-19 during 2020 and 2021. We utilized recent population data provided by the Program of Data Analysis for Research and Innovation in Health (PADRIS[8]). By comparing both datasets, we quantified changes over the past decade and estimated the associated uncertainty. The overall population increased by 2.5%. The correlation of population per ABS is very strong, as indicated by a Pearson's correlation coefficient of 0.97 (see Figure S.1 for the entire Catalonia). Detailed data for the four provinces are presented in Table S.2. The Root Mean Square deviation is approximately 10%.

##### 1.2 Temporary workers in the agricultural sector

During the summer season, Catalonia experiences an influx of temporary workers in the agricultural sector. The Statistical Institute of Catalonia, IDESCAT[13], provides data on the size and distribution of farming companies

**Table S.1.** Number of individuals per age category and some relevant groups, as implemented in the simulation.

| Age (years) | Individuals (#) |
|---|---|
| < 15 | 1,182,921 |
| 15 – 64 | 4,987,483 |
| > 64 | 1,244,494 |
| Nursing facilities residents | 53,782 |
| Nursing facilities workers | 33,952 |
| Hospital workers | 28,270 |
| Temporary workers | 4,143 |
| Total | 7,535,045 |

**Table S.2.** Comparison between the number of people per ABS as provided by the healthcare system and the census data (not including temporary workers).

| Area | Census | Healthcare System | Increase in % | Pearson's correlation coefficient |
|---|---|---|---|---|
| Catalonia | 7,530,902 | 7,722,113 | 2.5% | 0.97 |
| Barcelona | 5,446,555 | 5,615,924 | 3.1% | 0.96 |
| Girona | 851,123 | 873,990 | 2.7% | 0.99 |
| Tarragona | 791,469 | 796,356 | 0.6% | 0.99 |
| Lleida | 441,755 | 435,843 | 2.6% | 0.99 |

based on information from the "Directori central d'empreses" (DIRCE)[14]. This data includes details about worker composition and the count of temporary workers. Typically, Catalonia receives approximately 6,500 temporary workers during the summer, with the province of Lleida hosting the largest proportion. We created an additional segment of the census file with 4,143 temporary workers concentrated in areas with higher agricultural activity, distributed across the counties of Segrià, Pla d'Urgell, Urgell, Noguera, Alt Empordà, Alt Penedès. The workers are randomly assigned to mock farming companies in these regions, with the number of workers per company generated according to a Poisson distribution (20 workers for fruit picking in the first four regions and 5 workers for grape harvesting in vineyards in the latter two). In our simulation, we assume that the age profile of these workers ranges from 18 to 44 years old. Based on local statistics, we assume that these workers reside in the same ABS as their workplace and share housing. They are assigned social contacts like the rest of the population. The temporary workers are progressively introduced into the population during the first two weeks of July of 2020. The same casuistic was not repeated in summer 2021 in the simulation, as data did not show signs of the same level of contagion likely due to increased protection measures.

##### 1.3 Mobility data

In this study, we use two sets of processed mobility data sourced from the INE[28] and from the Barcelona Supercomputing Center (BSC)[10]. The ratio between the average daily mobility derived from the BSC and INE datasets is approximately 1 for work-related activities but close to 2 for leisure activities, as shown in Figure S.2. During outbreak peaks, this ratio tends to increase, indicating a more significant reduction in longer trips (as selected by INE) compared to shorter trips (as selected by BSC).

A calibration factor, which depends on the level of restrictions for leisure activities, translates the change in mobility into an estimate of the reduction in effective viral load (see Table S.8). Detailed mobility information from

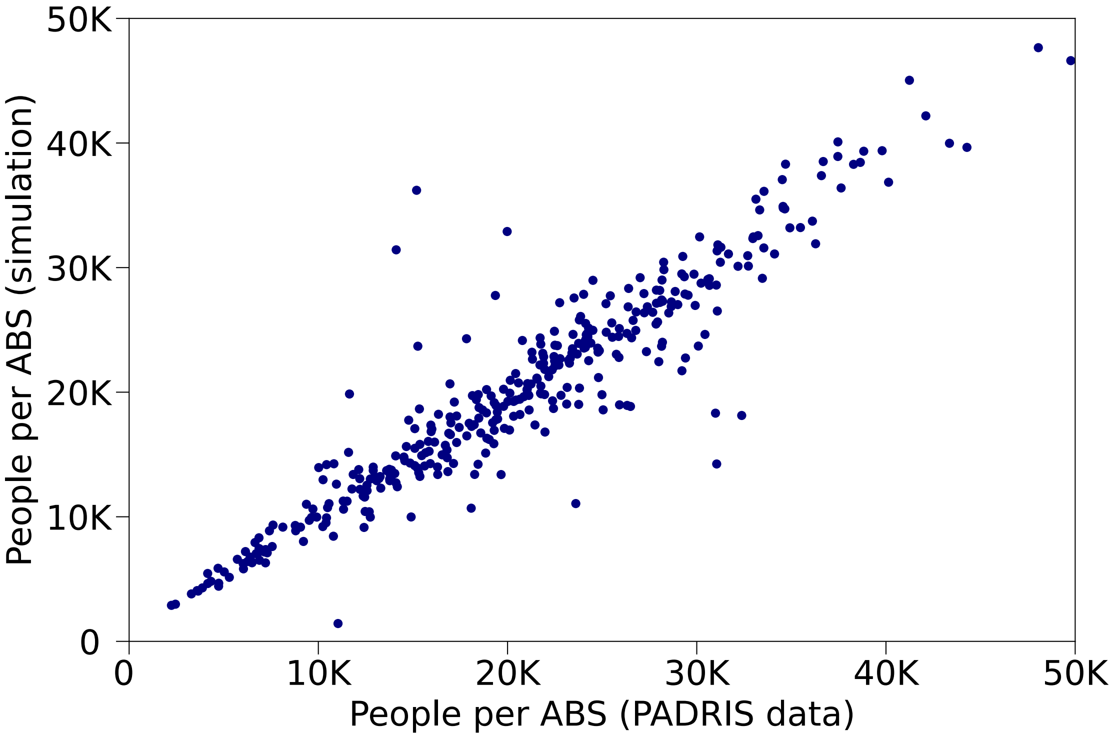


**Figure S.1. Population per ABS.** Simulated population per ABS according to the 2011 census data vs the 2020 Catalan healthcare system data.

BSC was only available until week 62. For the remainder of the period, we relied on the average INE data to adjust mobility, maintaining the last-day relative ratios between ABSs.

#### 2 Modelling of the spread of the disease

##### 2.1 Calculation of the probability of infection

The probability of infection $P$ is given by the expression (1) of the main document, expression that we report here for convenience,

$$P = 1 - e^{-\lambda_i \cdot F^i_{\text{EfficiencyVaccine}}(t) \cdot \Delta t}. \tag{S.1}$$

We calculate the viral load exposition $\lambda_i$ of Equation S.1 during the three eight-hour intervals that an individual $i$ spends either at home (Home), at work or school (Work), or in social interactions (Leisure). In addition, individuals may use public transport (PT) to go to work or in the context of social activities (WorkPT or LeisurePT). Additional interactions may take place during the summer in crowded settings (SummerActivities). The force of infection $\lambda$, where $\lambda$ represents a vector of 7.5M entries, one per individual, is thus expressed as a sum of terms:

$$\lambda(t) = \lambda_{\text{Home}}(t) + \lambda_{\text{Work}}(t) + \lambda_{\text{Leisure}}(t) + \lambda_{\text{WorkPT}}(t) + \lambda_{\text{LeisurePT}}(t) + \lambda_{\text{SummerActivities}}(t)\,. \tag{S.2}$$

Depending on the time interval, one or more terms may contribute. To calculate $\lambda$, we first have to quantify the viral shedding $\kappa$ of every infectious individual. It results from the product of several factors:

$$\begin{aligned}\kappa^i(t) =& I^i_{\text{Infectiousness}} \times F^i_{\text{Vaccine}}(t) \times F^i_{\text{Reinfected}}(t) \times F^i_{\text{Residency}} \times F^i_{\text{TimeProfile}}(t - t^i_0) \times F_{\text{Contagiousness}} \times \\ & \times F_{\text{Mask}}(t) \times F^i_{\text{MaskWearing}}(t)\,.\end{aligned} \tag{S.3}$$

The first factor $I^i_{\text{Infectiousness}}$ represents the overall strength of viral load shedding of an individual, the second $F^i_{\text{Vaccine}}(t)$ the reduction in infectiousness intensity resulting from the vaccine, the third $F^i_{\text{Reinfected}}(t)$ the reduction

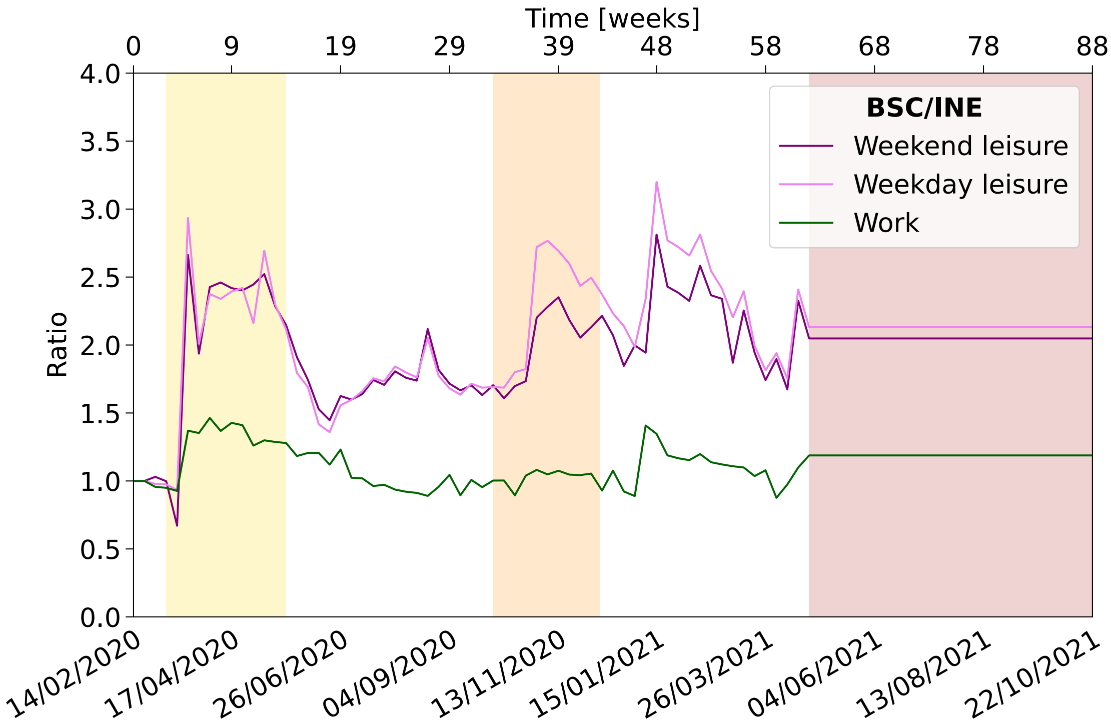


**Figure S.2. Mobility Evolution Ratio of BSC over INE.** The ratio compares the average daily mobility data for work/school and leisure activities between BSC and INE in Catalonia. The analysis covers weekdays and weekends, highlighting differences in mobility. Shaded vertical bands highlight periods of varying mobility restrictions, corresponding to the first, second, and fourth/fifth waves of the pandemic.

in infectiousness for reinfected people, the fourth $F^i_{\text{Residency}}(t)$ the reduction in transmission for residents of large nursing homes, the fifth $F^i_{\text{TimeProfile}}(t-t^i_0)$ the relative amount of viral shedding as a function of the time elapsed since the onset of infectiousness $t^i_0$, the sixth $F_{\text{Contagiousness}}$ an overall calibration factor that converts viral load into the corresponding force of infection, the seventh $F_{\text{Mask}}(t)$ the reduction in viral transmission provided by face masks, based on the enforcement policy at that time, and the eighth $F^i_{\text{MaskWearing}}(t)$ the effectiveness with which individuals wear their mask. Next, we evaluate the contacts of every individual during that time interval. We distinguish two types of contacts[2]: those occurring within networks of individuals, such as people sharing the same home or workplace, and incidental contacts, such as those occurring on public transport. The first three terms in Equation S.2 pertain to the first type and are described by contact matrices $A$ of dimension (7.5M $\times$ 7.5M), as detailed in the next section. The vector $\lambda$ results from the product $\lambda = A \times \kappa$, where $\kappa$ is a vector of dimension 7.5M. For incidental contacts, we do not identify the specific individuals involved but instead calculate the average viral shedding $\overline{\kappa}$ of the group of people potentially involved, e.g. all individuals using public transport to commute to work. The number of contacts is given by a Poisson variation of an estimated mean number of contacts (MC), such that $\lambda^i = \text{Poisson}(MC)^i \times \overline{\kappa}$.

#### 2.2 Matrices to model contacts at home, work/school or and leisure

The contact matrices are constructed differently for the three environments. For home contacts, we use a simple model where all contacts between residents are equally probable, with an average home size of 2.73 individuals. In nursing homes, the number of residents can be quite large, sometimes up to 200, which would theoretically lead to a much faster progression of contagion than what is observed in reality. To correct for this, we apply an empirical factor to the viral load emission of residents $1/(\text{number of residents})^N$ to reduce the effective duration of the contacts between individuals, as previously suggested[29].

Regarding work contacts modelling, in our initial study, we only considered contacts within classrooms or subgroups within companies. To more accurately reflect the size of larger companies, we implemented a contact model where 80% of contacts occur within subgroups and 20% within the larger groups. The number of contacts within classrooms is ten, while it is seven within company subgroups. We use the NetworkX[30] package with the Havel-Hakimi algorithm[31] to model contacts within the subgroups. This approach provides heterogeneous degree distributions and an increasingly higher clustering, which are both crucial features of complex, real networks. See Table S.7.

For the stable leisure contacts, each individual is assigned a specific ABS for weekday leisure activities, and potentially a different one for the weekend, as indicated by the mobility data. Groups of friends are selected from the inhabitants of these ABSs, with the group size determined by a Poisson random fluctuation around an age-dependent average, typically around five, derived from the most recent and comprehensive synthetic contact matrices set[33]. The list of leisure ABSs changes for the fraction of the population that relocates during the summer, according to corresponding mobility data. We use the NetworkX Configuration model[32] to randomly select contacts within the corresponding ABS, without age selection.

For incidental contacts, we use a different approach. We define an average number of contacts occurring during these activities and calculate the average viral load of the population segment involved, such as people using public transport or those in summer leisure ABSs. This method allows us to consider the impact of tourism, with 500k visitors from outside Catalonia during the summer of 2020 and approximately double in 2021[13]. We assume that the average infection rate among tourists is the same as the local population. The impact is particularly noticeable in tourist areas, primarily along the coast and in central Barcelona. These additional contacts are incorporated during the summer periods and other holidays, such as Easter in April and Christmas in December.

Certain categories of individuals are treated separately to account for specific circumstances. To consider the contacts between nursing sanitary workers and residents of nursing homes, we assume that they interact during an 8-hour working time slot. Both residents and workers wear masks as required by authorities. For hospital workers, we assume that 65% of them are in direct contact with an average of four hospitalized COVID-19 patients. This percentage corresponds to the number of vaccinated workers at the beginning of the vaccination process. We introduce a modulation factor, the relative mobility per ABS, to accommodate local variations in hospital occupancy.

#### 3 Parameters setting and calibration procedure

The proposed model relies on 194 parameters to characterize the population, the pattern of contacts, the evolution of the disease, the lockdown and self-protection measures and the vaccination campaign. We initially set their values according to our best knowledge based on external information as described in a previous publication[2] and further input (see Section 1). In addition, we use subsets of PADRIS[8] data governing specific details of the modelling to set the corresponding parameters (e.g. the ABS where the first flares of diagnosed cases occurred to define the location of some of the initially infectious people, or the number of sanitary workers diagnosed in geriatrics and hospitals to set the average number of worker contacts with residents or patients). Details are given in Section 3.1.

On the other hand, some parameters have a more global impact affecting various age categories of the population across the full-time interval under study and in the full territory (a good example is the overall strength of the viral load emitted by infectious individuals). We identified these parameters and performed a goodness-of-fit test to further calibrate the model as discussed in Section 3.2.

All parameter values are listed in Tables S.6 to S.13.

##### 3.1 Parameters settings

Many parameters of the model were already defined in our initial study of the province of Barcelona[2]. In this section, we focus on the improvements and the complementary information needed to extend the model to the entire Catalan territory.

The simulation starts on 14 February 2020. We assume that a small initial randomly selected fraction of the population (0.0015% or about 70 people) was already infected. The analysis of data from the initial period revealed that 10 ABSs experienced early outbreaks. We then added two randomly chosen individuals in each of these ABSs to the initially infected set, while excluding those from the Lleida province due to the slower progression of the disease in that region. Further details are provided in Tables S.6 and S.11.

The amount of time individuals spend at home, work, or engaging in leisure activities varies over time and depends on their category. Typically, an adult worker spends eight hours at home, eight hours at work during the week, and eight hours in leisure activities. This pattern also applies to school pupils and university students. During weekends, when work or school is not in session, people allocate their time to leisure activities. Individuals who are not employed are assumed to remain at home during the week. School and university pupils are categorized as non-workers during holiday periods. Additional categories are defined to reflect situations such as confinement, where individuals stay at home instead of going to work and reduce leisure activities (see Table S.7). Moreover, to reflect the reality of teleworking during confinement, a fraction of workers stay at home instead of commuting to work. This fraction is determined based on the relative evolution of work mobility over time.

The set of parameters that control the model of contacts described in Sections 2.2 are provided in Tables S.7 and S.11. The parameter governing the reduction of the effectiveness of contacts between residents in large nursing homes was set to 0.4, according to data from the initial wave.

The amount of incidental contacts and their time profile depend on the region, especially during the summer. We analysed the data of summer 2020 with the granularity of the 32 "Sectors Sanitaris"[13] and defined different levels of contacts, ranging from 1 to 3, as well as their time onset with Barcelona featuring the earliest one. Summer 2021 is assumed to be similar but with a factor of 2.5 more contacts, reflecting the increase of influx of tourists[13]. Table S.7 gives the details.

The comparison of the mobility of INE and BSC (see Section 1.3 and Figure S.2) shows differences in their relative level that are more pronounced during the strong confinement periods. We implemented the corresponding normalisation factors in the simulation as described in Table S.8.

The characteristics of the disease evolution are detailed in Table S.6, and the effectiveness of mask-wearing in Table S.12.

The probability of being diagnosed depended initially mostly on the severity of the symptoms. Later more tests were made on people that were in contact with diagnosed people, or preventively before holiday periods. We used the PADRIS data[8] of the total number of tests performed as a function of time and age to model the increase of tests during the waves. Table S.8 provides the parameters.

The parameters of the model reproducing the age and time-dependent pattern of vaccination data shown in the main paper are given in Tables S.8 and S.13.

##### 3.2 Parameter Fitting procedure

Firstly, it is important to note that direct comparison between the daily results produced by the model and real-life collected data is challenging due to the observed weekly dips in registered cases, as depicted in Figure 2 of the main paper. These dips can be attributed to a combination of factors, including fewer staff conducting tests on patients during the weekend and a lower likelihood of patients seeking medical attention during this time. To address this issue, we aggregate all cases on a weekly basis, with weeks considered from Friday to Thursday. This choice is motivated by the assumption that many cases not recorded on a specific day are eventually captured in the count over the following days. If we were to aggregate data on a Monday to Sunday week, it could result in undercounting cases during periods of increasing incidence and overcounting them during periods of decreasing incidence. As a result, all waves would appear to be delayed by a few days.

Additionally, performing a direct multi-parametric fit on the model would not only be highly computationally demanding but also conceptually unfeasible since there is no precise understanding of the ranges of variation and the correlations of all the parameters. We use instead a single parameter fit to study several of the more relevant parameters. As such, we make several simplifications in order to streamline the parameter fitting process:

1. Firstly, we fit one parameter at a time while keeping all other parameters fixed. The result is then used to study the next parameter, continuing this process until all desired parameters are fitted. The sequence in which parameters are fitted is determined based on the sensitivity of the simulations to small changes in the parameter value (see Section 3.3).

2. Secondly, rather than executing the simulation code at each iterative step in the fitting process, we pre-generate a discrete set of simulations using various values of the parameter under study. We then interpolate between these simulations to generate our "model". This approach significantly reduces the computational cost of the algorithm while maintaining accuracy to a reasonable extent.

3. Finally, only the first year of the simulations is taken into account, given that the introduction of vaccines heavily modifies the behaviour of all the relevant data.

The cost function employed in order to fit the model to real data is based on a $\chi^2$ statistic. In addition to the purely statistical Poisson uncertainty of the data, we consider the following source of uncertainties in the fit:

1. An uncertainty due to the stochastic nature of the processes involved in the generation of every simulation. This uncertainty was quantified by generating 51 random samples varying the 26 different seeds and calculating the relative standard deviation of the resulting simulations.

2. A systematic uncertainty due to discrepancies during the data-collecting process. Several data sources may report different numbers of registered patients each day during the pandemic, depending on the criteria used for their selection. Upon comparing these sources, the relative uncertainty remains within an envelope of approximately 10%.

As discussed above, accurately quantifying all systematic uncertainties related to the imperfect knowledge of many parameters is challenging. Therefore, the uncertainties considered in the fit should be viewed as a lower limit.

Thus, starting from an initial value for the chosen parameter, the fitting algorithm generates a test simulation by interpolating between the closest pre-generated ones available and performing a goodness-of-fit assessment based on its deviation from the collected data, while taking into account the uncertainties as described above. This comparison is performed for all possible combinations of province and age group, whose partial errors are then added up in quadrature to produce a single error measurement.

An additional consideration arises when dealing with smaller age groups and provinces. In some weeks, there may be a very small number of registered cases, or even none at all, which can lead to divergences in the fitting algorithm. To ensure reliable error calculation for any particular simulation, we only consider weeks with at least 10 registered cases. Figure S.3 shows the category with the highest statistical power, the adult population in Barcelona province, and Figure S.4 an example of the less numerous category, nursing facilities workers in the Tarragona province.

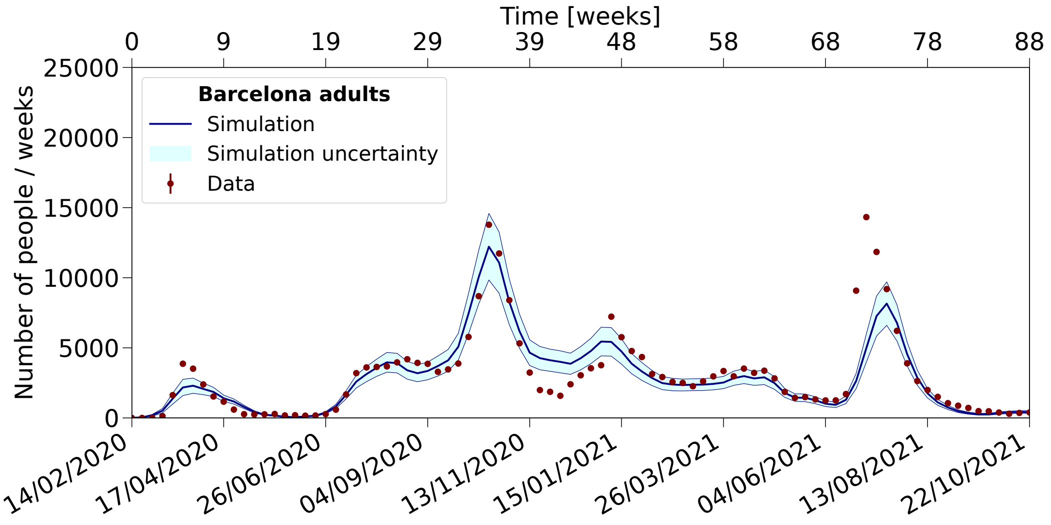


**Figure S.3. Number of diagnosed adults in the province of Barcelona.** Data on diagnosed adults in the province of Barcelona are compared to simulation results for the period from 2020 to 2021.

##### 3.3 Choice and fit of the most sensitive parameters

To gauge the sensitivity of a simulation to a specific parameter, we consider the relative variation in the total number of patients diagnosed during the first and second waves of the virus across Catalonia, as well as the maximum number of diagnosed cases on a weekly basis ("peaks"), when the parameter under study is modified by a $\pm 10\%$ from its central value. Given its much greater relative weight in the total number of cases for the first year of the pandemic, the second wave is given priority over the first one in terms of sensitivity estimations. Table S.3 presents all these quantities for the three most sensitive parameters. These three parameters were ultimately used in the calibration process, in the order they appear in the Table. The first parameter translates the "leisure" relative mobility variation to the reduction of viral load exposure during leisure activities (*MobiLeisure*), as discussed in Section "Mobility" of the main paper. The second parameter is the overall normalisation of the emitted viral load (*NormViral*) by a person affected by COVID-19. The third is a parameter that controls the relative distribution of people in the three different categories of infectiousness according to their age (*CatInfectious*), as discussed in Section "Model design for the COVID-19 spread" of the main paper.

#### 4 Predicted level of diagnosed people aggregated by categories

In this section, we present the number of diagnosed individuals aggregated in various age, professional and territorial categories.

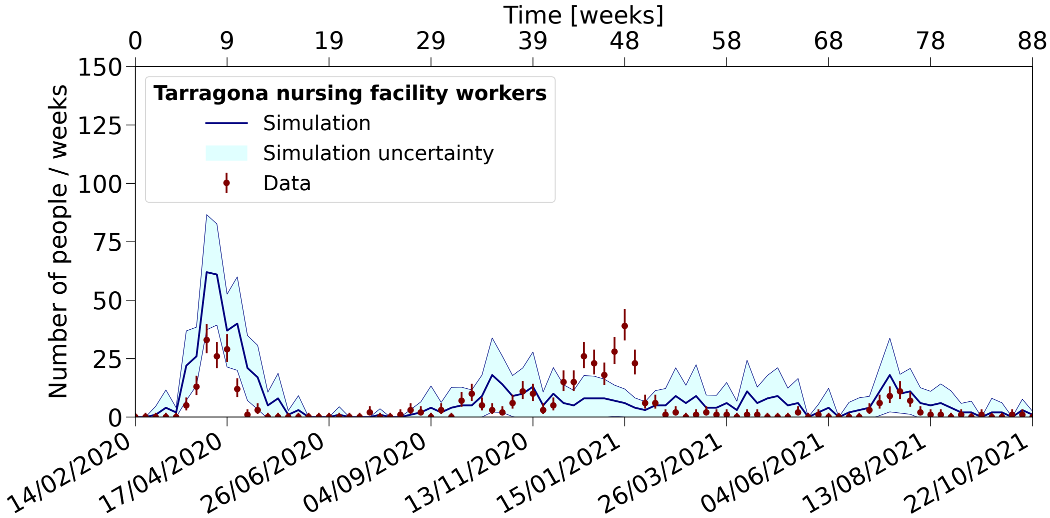


**Figure S.4. Number of diagnosed nursing facilities workers in the province of Tarragona.** Data on diagnosed nursing facilities workers in the province of Tarragona are compared to simulation results for the period from 2020 to 2021.

**Table S.3.** Maximum number of weekly diagnosed cases ("peaks") and total number of cases, along with their respective relative deviations, of the first two COVID waves across all provinces and age groups for the three most sensitive parameters.

| Parameter | | $1^{st}$ wave peak | $1^{st}$ wave total | $2^{nd}$ wave peak | $2^{nd}$ wave total |
|---|---|---|---|---|---|
| MobiLeisure | +10% | 7045 | 44924 | 86411 | 669260 |
| | -10% | 5453 | 36139 | 2193 | 35511 |
| | Relat. dev. | 0.25 | 0.22 | 1.90 | 1.80 |
| NormViral | +10% | 10310 | 66222 | 49627 | 438722 |
| | -10% | 4229 | 27343 | 2882 | 41339 |
| | Relat. dev. | 0.84 | 0.83 | 1.78 | 1.66 |
| CatInfectious | +10% | 9240 | 58109 | 28111 | 267581 |
| | -10% | 5226 | 34306 | 7755 | 85756 |
| | Relat. dev. | 0.55 | 0.52 | 1.13 | 1.03 |

#### 4.1 Wave patterns for different age groups of population

Figures S.5, S.6, S.7 show the weekly number of diagnosed individuals across three age categories: *Children, Adult* and *Senior*. The Adult category exhibits the best agreement, as expected due to its larger statistical sample size. The distribution features closely resemble those of the total population, as illustrated and discussed in Figure 6 in the main paper. The main differences in the shape of the data are seen in the case of the initial wave. Children are mostly asymptomatic, seniors tend to display stronger symptoms and higher infectiousness. During the early stages of the pandemic, testing primarily targeted individuals with severe symptoms, resulting in fewer diagnosed cases among children and relatively more among seniors compared to adults. The simulation effectively reproduces these distinctions. Later on, a broader spectrum of people were tested, including close contacts of diagnosed people. The simulation tends to overestimate the height of the summer plateau in simulation compared to data in the senior category and to a lesser extent in the case of children. This is likely related to leisure contacts, the relevant ingredient in that case as seen in Figure S.15. The pattern of contacts is assumed to be the same for the different age categories and may be overestimated in the case of seniors and children. This may also explain the excess observed in the simulation for these categories during the summer of 2021.

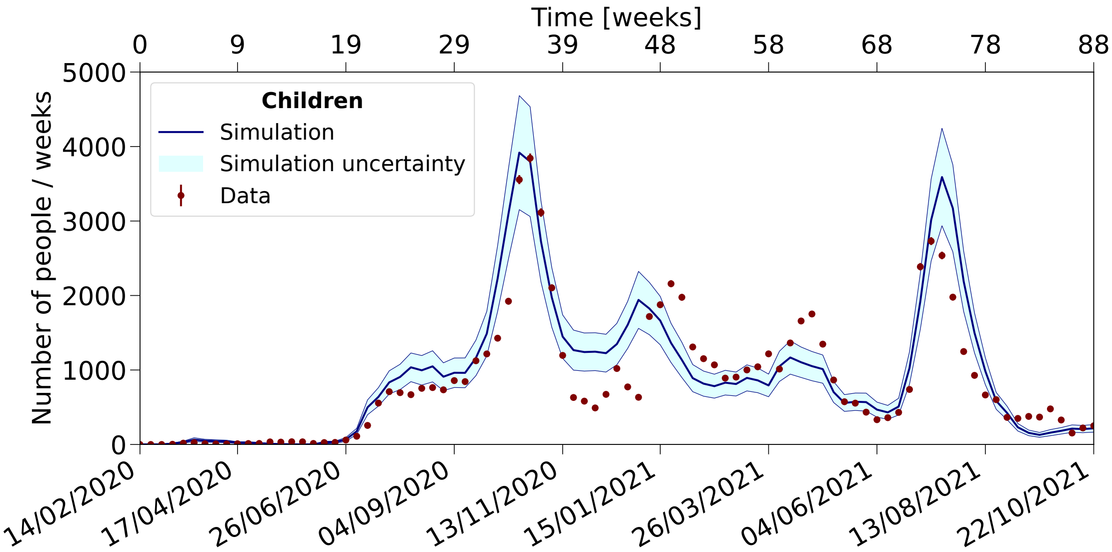


**Figure S.5. Number of diagnosed children in Catalonia.** Data on diagnosed children are compared to simulation results for the period from 2020 to 2021.

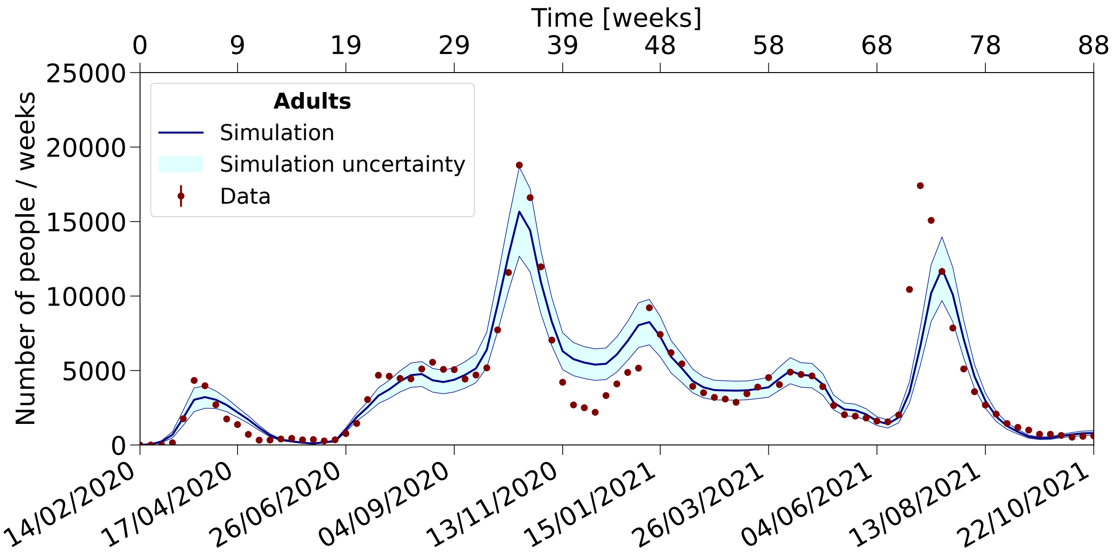


**Figure S.6. Number of diagnosed adults in Catalonia.** Data on diagnosed adults are compared to simulation results for the period from 2020 to 2021.

#### 4.2 Wave patterns for nursing facilities and sanitary workers

Figures S.8, S.9, S.10 show the data for nursing facilities residents, nursing facilities sanitary workers and hospital workers (sanitary workers in contacts with infectious people in hospitals). The first wave is quite well reproduced for nursing facilities residents and both categories of sanitary workers. The October and December waves are somewhat underestimated. One of the important factors affecting these waves are leisure contacts, which may be underestimated in the residencies.

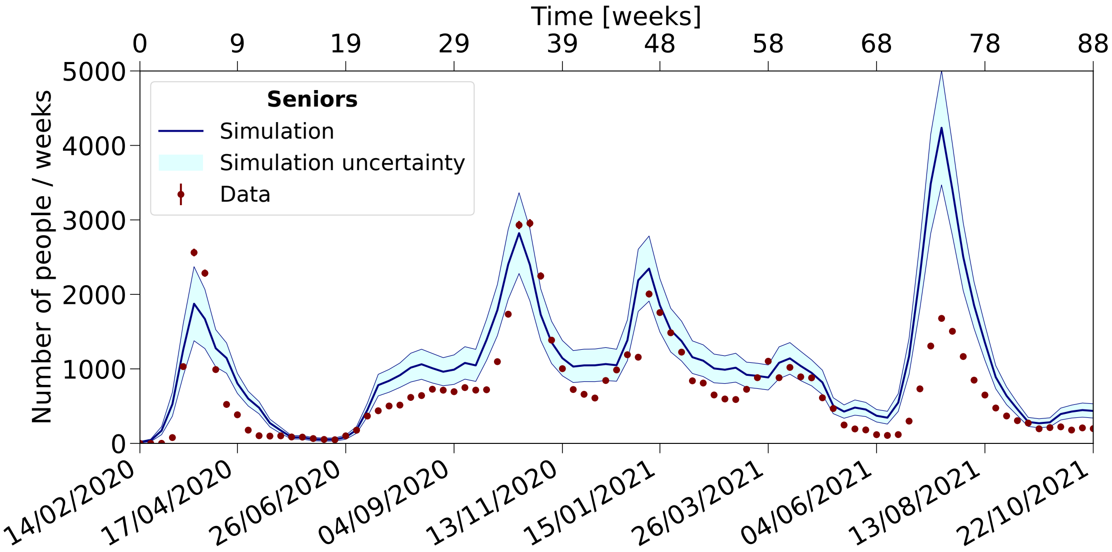


**Figure S.7. Number of diagnosed seniors in Catalonia.** Data on diagnosed seniors are compared to simulation results for the period from 2020 to 2021..

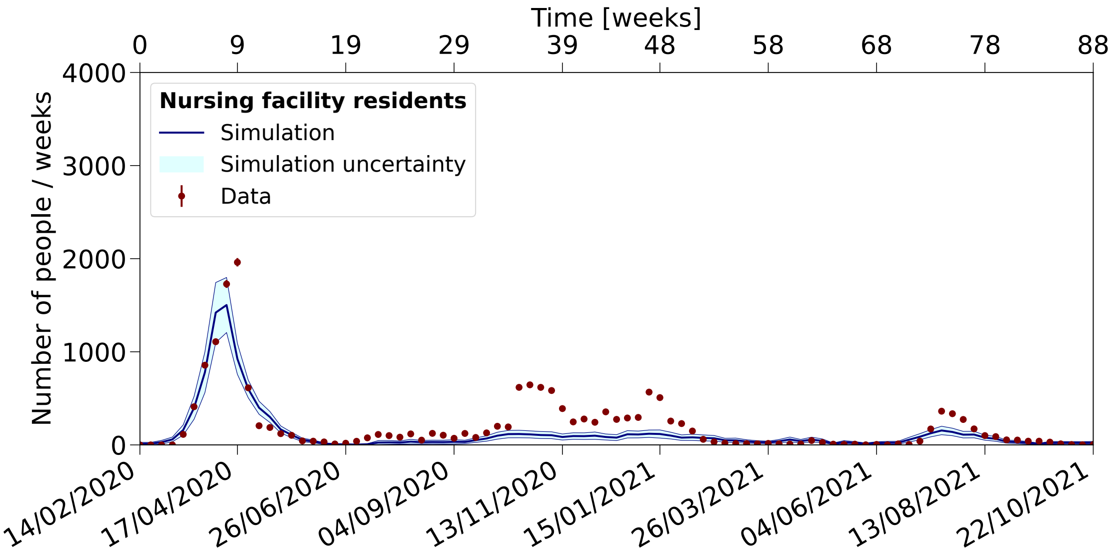


**Figure S.8. Number of diagnosed nursing facilities residents in Catalonia.** Data on diagnosed nursing facilities residents are compared to simulation results for the period from 2020 to 2021.

##### 4.3 Wave patterns for the four provinces and their correlation

Figures S.11, S.12, S.13, S.14 show the data for the Barcelona, Girona, Lleida and Tarragona provinces. The features of the Barcelona province are most accurately reproduced, given its significantly larger population and greater statistical power. When comparing different provinces, it is relevant to underline that we use the mobility information at the level of ABS. The three outer provinces tend to have shallower minima of mobility compared to Barcelona during confinement periods and to recover a higher mobility. Variations are typically of the order of 10%. Such differences have a substantial impact on the propagation of the virus, underscoring the importance of using high-granularity spatial information, including the movement of the population leaving Barcelona during the summer. Using mobility data averaged over all provinces instead of individual ABS information can lead to variations as large as a factor of two in predictions, significantly compromising the observed level of agreement.

The first wave tends to be overestimated in the three outer provinces, as well as in some of the later waves.

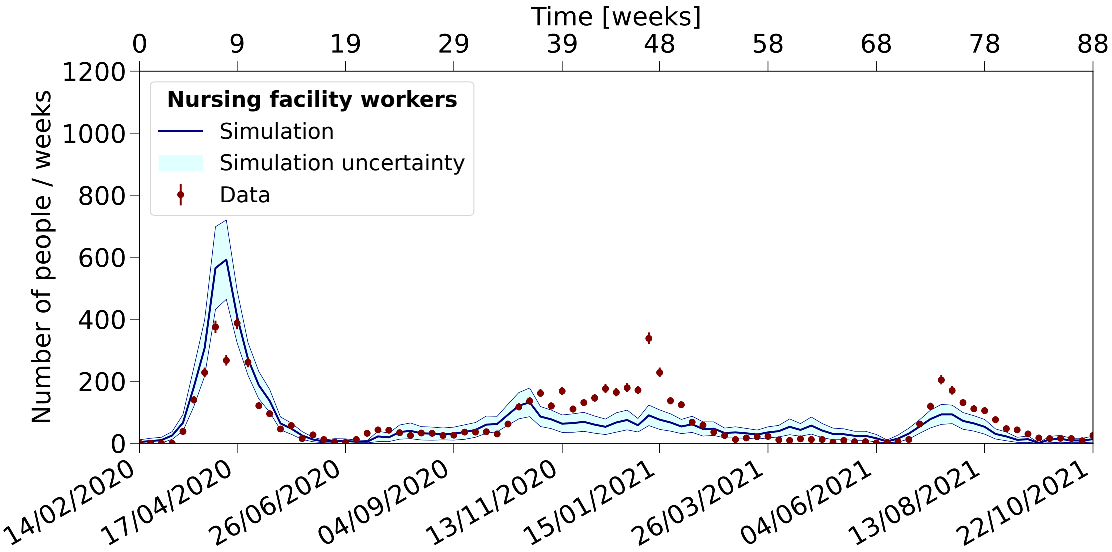


**Figure S.9. Number of diagnosed nursing facilities sanitary workers in Catalonia.** Data on diagnosed nursing facilities sanitary workers are compared to simulation results for the period from 2020 to 2021.

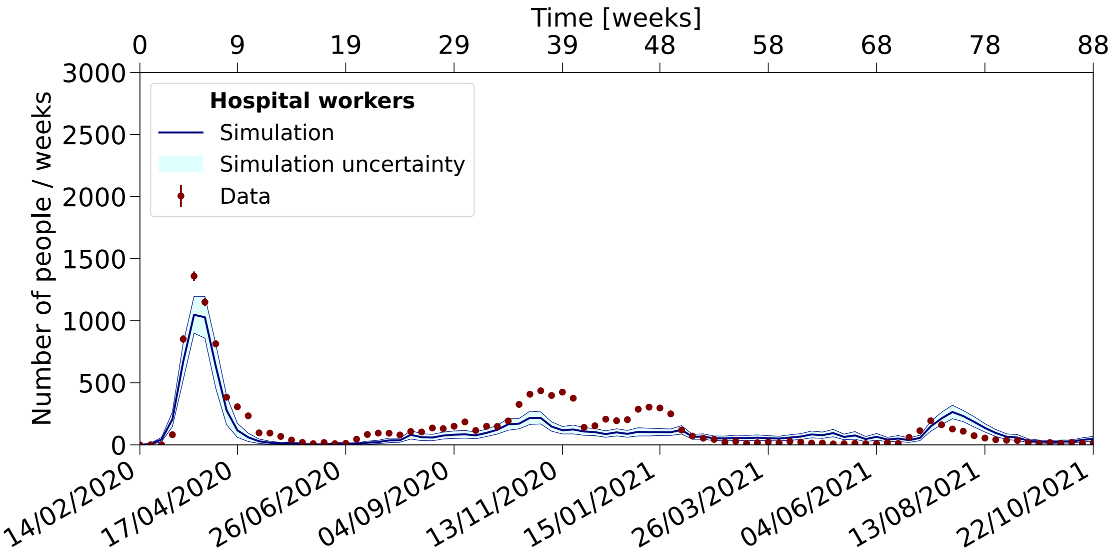


**Figure S.10. Number of diagnosed hospital workers in Catalonia.** Data on diagnosed hospital workers are compared to simulation results for the period from 2020 to 2021.

This discrepancy is likely due to differences in the pattern of leisure contacts, which may vary in these less densely populated areas compared to Barcelona. It is worth noting the unique case of the Lleida province, where a distinct wave is observed in July 2020, coinciding with the arrival of temporary agricultural workers.

We examined the correlation between the daily evolution of waves across different provinces to gain insight into their nature, as previously discussed[24]. Table S.4 shows the Pearson's correlation coefficients between diagnosed cases in the four provinces during the March and October 2020 waves, as well as the summer period, for both observed data and simulation results. The correlation is high during the two waves, reflecting the synchronous spread of the virus, while it is lower during the summer, with more holiday activities and foreign tourists in certain areas. Notably, in cases involving the Lleida province, strongly affected by the influx of temporary agricultural workers, the correlation coefficients may even be negative.

Table S.5 shows the level of contacts across provincial borders during leisure activities, as extracted from the

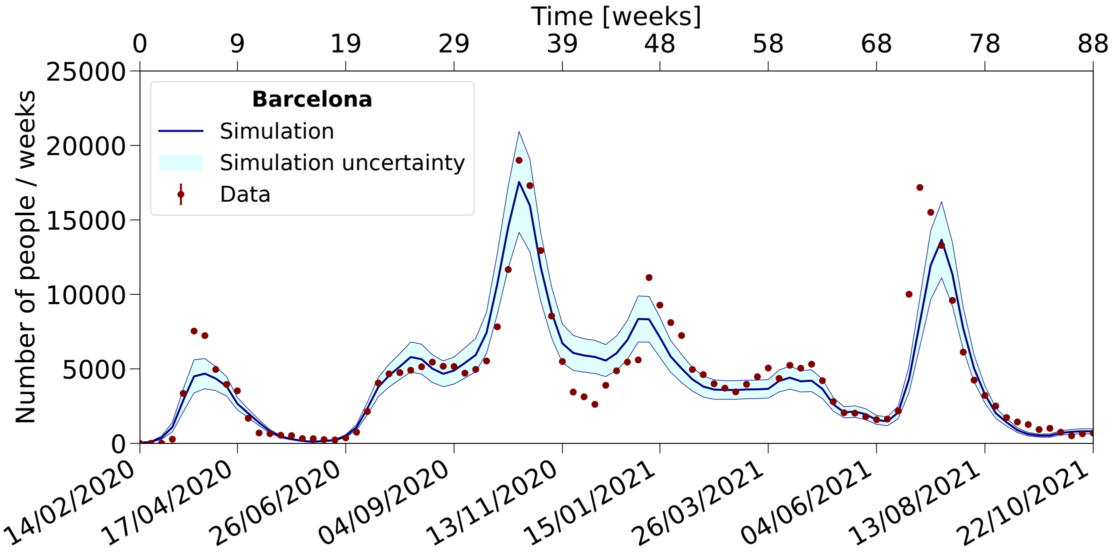


**Figure S.11. Number of diagnosed people in the province of Barcelona.** Data on diagnosed people are compared to simulation results for the period from 2020 to 2021.

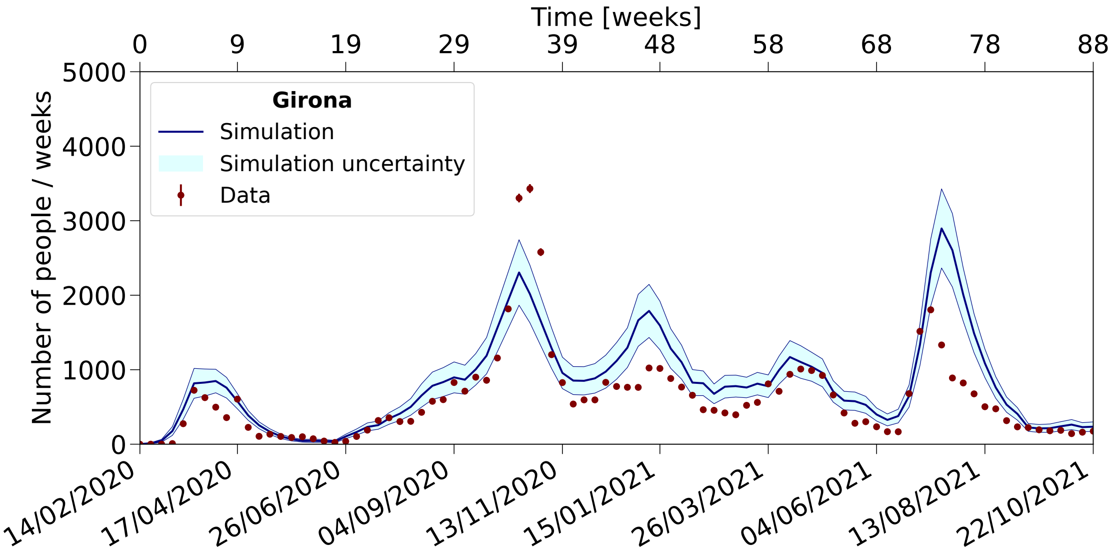


**Figure S.12. Number of diagnosed people in the province of Girona.** Data on diagnosed people are compared to simulation results for the period from 2020 to 2021.

mobility data. It reveals higher exchange rates between Barcelona and its neighbours that affect more strongly less populated provinces, with significant impacts during summer as residents from Barcelona (up to about 400 thousand people) travel to Mediterranean coastal resorts and the Pyrenees region. The relative impact in the provinces of Girona, Tarragona, and Lleida is 30, 20, and 10%, respectively. This effect is incorporated into the simulation, reallocating part of the population to different ABSs during summer and adjusting the list of social contacts accordingly.

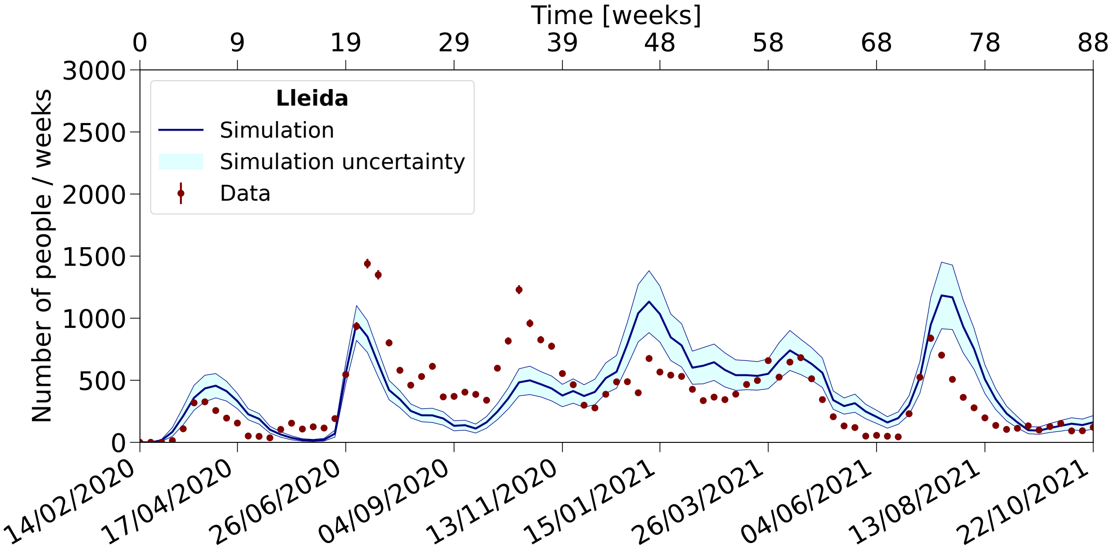


**Figure S.13. Number of diagnosed people in the province of Lleida.** Data on diagnosed people are compared to simulation results for the period from 2020 to 2021.

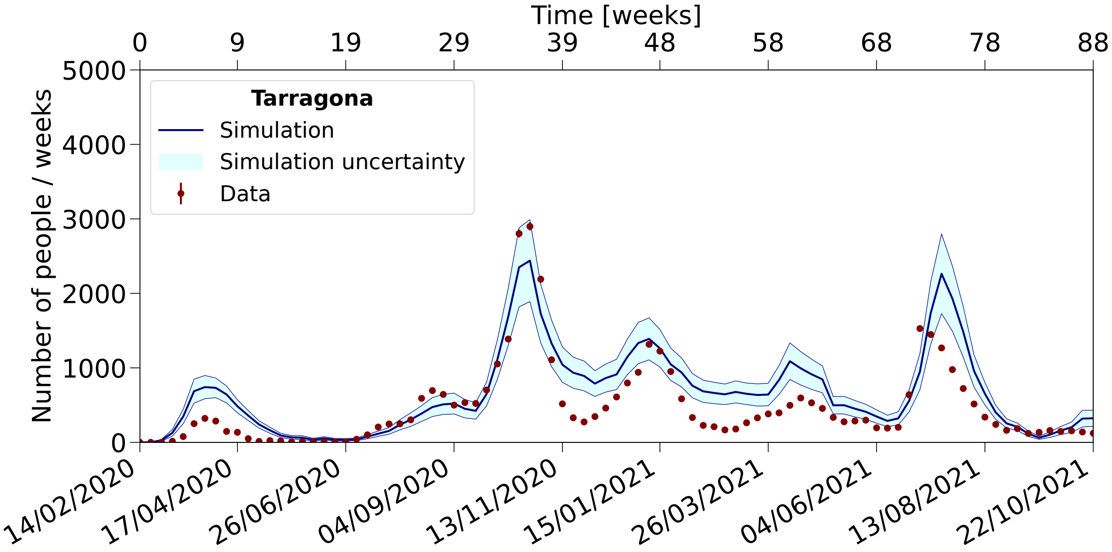


**Figure S.14. Number of diagnosed people in the province of Tarragona.** Data on diagnosed people are compared to simulation results for the period from 2020 to 2021.

**Table S.4.** Pearson's correlation coefficient between the number of people diagnosed by PCR tests between the province of Barcelona and the other provinces during different periods of 2020.

| Wave | Data type | Bcn-Gir | Bcn-Tar | Bcn-Lle | Tar-Gir | Tar-Lle | Gir-Lle |
|---|---|---|---|---|---|---|---|
| March | Simulation | 0.96 | 0.90 | 0.82 | 0.93 | 0.86 | 0.87 |
| | Data | 0.79 | 0.88 | 0.88 | 0.75 | 0.82 | 0.72 |
| Summer | Simulation | 0.69 | 0.72 | -0.93 | 0.94 | -0.82 | -0.81 |
| | Data | 0.82 | 0.64 | -0.13 | 0.80 | -0.35 | -0.06 |
| October | Simulation | 0.95 | 0.90 | 0.54 | 0.82 | 0.76 | 0.48 |
| | Data | 0.96 | 0.87 | 0.88 | 0.88 | 0.75 | 0.81 |

**Table S.5.** Population exchange between provinces in leisure activities, during the year and during the summer period, normalized to the total number of inhabitants of each province.

| Provinces | | Usual during the year | | Specific to summer | |
|---|---|---|---|---|---|
| Prov.1 | Prov.2 | in % of Prov.1 | in % of Prov.2 | in % of Prov.1 | in % of Prov.2 |
| Barcelona | Girona | 0.7 | 4.6 | 5.0 | 31.8 |
| Barcelona | Tarragona | 0.4 | 2.9 | 3.0 | 20.4 |
| Barcelona | Lleida | 0.4 | 4.4 | 0.8 | 10.4 |
| Girona | Tarragona | 0.0 | 0.0 | 0.0 | 0.0 |
| Girona | Lleida | 0.6 | 1.2 | 0.8 | 1.5 |
| Tarragona | Lleida | 0.1 | 0.3 | 0.3 | 0.6 |

### 5 Components of viral load

We examine the relative importance of the various components of the total viral load the population is exposed to during 2020 and 2021. Figure S.15 shows the three main categories of activities: at home, at work and during leisure activities. The latter is further divided into stable ("Leisure") and incidental contacts ("Extra leisure"). Initially, during the first 2-3 weeks, leisure activities dominate contagion. Subsequently, the importance of work-related contacts increases progressively, eventually giving way to dominance by home-based contacts, especially during the first wave with strict confinement measures (see top figure). During the summer, as the leisure activities resumed, the "Leisure" category regained importance, along with the incidental contacts captured by the "Extra leisure" category (see bottom figure).

In the simulation, we observe that the leisure categories during the summer play a crucial role in increasing the overall incidence level and precipitating the flare-up of the October wave. This is particularly true for "Extra leisure" as it facilitates disease spread beyond the bubbles of stable contacts. The contribution of "Work" after the first wave is somewhat limited, as pre-pandemic mobility levels were not fully restored, teleworking became more widely accepted, and mask usage increased. In 2021, the impact of the vaccination campaign was significant, as shown in Figure 7 in the main paper. Despite an increase in leisure contacts during the summer, the corresponding wave was effectively managed and kept under control.

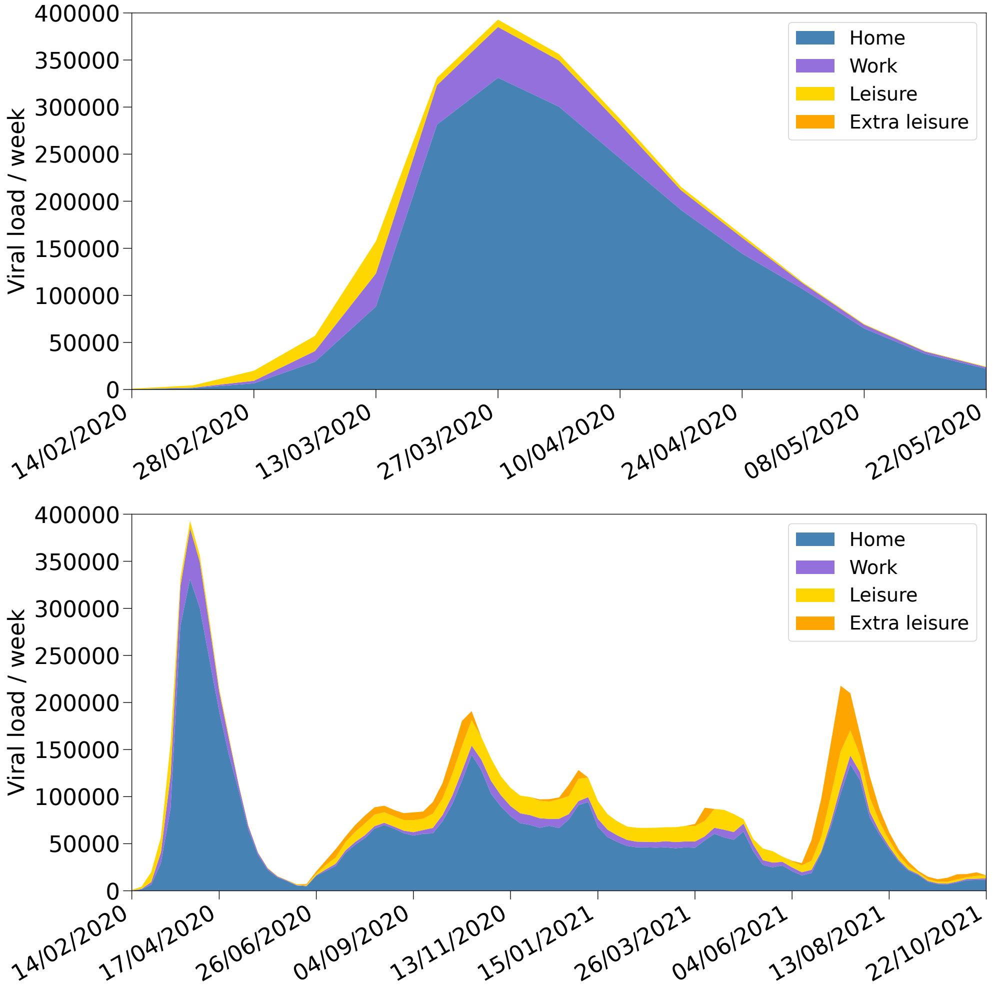


**Figure S.15. Components of the total viral load.** The total viral load to which the population is exposed is shown for the three main categories of activities: at home, at work, and during leisure activities, the latter distinguishes stable ("Leisure") and incidental contacts ("Extra leisure"). The top figure shows the first 15 weeks in 2020 and the bottom figures 89 weeks over 2020 and 2021.

#### 6 Tables with model parameter values

The model used to simulate the spread of COVID-19 includes a total of 194 parameters, which characterize the disease (53 parameters), contacts (82), lockdown and self-protection measures (19), and the vaccination campaign (40). Tables S.6 to S.12 describe the parameters, their settings and related references.

**Table S.6.** Parameters of the model (part Ia).

| Category | Number | Parameters | Source |
|---|---|---|---|
| | | Disease | |
| Incubation time (Gamma distribution) | 2 | $\mu, \sigma$ = 4.58, 3.24 days | Medical data[34,35,36] |
| Infectiousness time profile $F_{\text{TimeProfile}}$ (Gamma distribution) | 2 | $\mu, \sigma$ = 2.5, 1.7 days | Medical data[34,35,36] |
| Onset of infectiousness in relation to onset of symptoms | 1 | -2 days | Medical data[35] |
| Relative viral strength of asymptomatic, moderate and severely infectiousness | 3 | $I^i_{\text{Infectiousness}}$ = 0, 1, 2 | Medical data[36] |
| Viral strength to probability of infection conversion | 1 | $F_{\text{Contagiousness}}$ = 4.26 | 2$^{\text{nd}}$ fitted parameter |
| Fraction in categories of viral strength (age dependent) | 6 | See table S.9 | Medical data[36], 3$^{\text{rd}}$ fitted parameter |
| Reduction of infectiousness in case of reinfection | 1 | 0.8 | Assumed + Medical data[36] |
| Probability to be diagnosed in 1$^{\text{st}}$, 2$^{\text{nd}}$ & 3$^{\text{rd}}$, 4$^{\text{th}}$, 5$^{\text{th}}$ waves, dates & width | 17 | $P_D^{ANI}$ = (0.0,0.4,0.7,0.5) $P_D^{AMI}$ = (0.0,0.5,0.7,0.7) $P_D^{SSI}$ = (1.0,1.0,1.0,1.0) 20/7/20, 4/11/20, 26/1/21, 5/8/21, 90 days | Medical data[37] and PADRIS data[8] |
| Probability to be hospitalized, UCI & death | 3 | 0.5, 0.15, 0.01 | Assumed + Medical data[37] |
| Diagnosis time (Poisson distribution) | 3 | $\mu$, min., max. = 6, 3, 14 days | Medical data[37] |
| Recovery time | 1 | 14 days | Medical data[38] |
| Initially exposed | 12 | 0.0015% + 2 pers. in 10 ABS | Assumed + PADRIS data[8] |

**Table S.7.** Parameters of the model (part Ib).

| Category | Number | Parameters | Source |
|---|---|---|---|
| | | Contact's model | |
| Classroom size (fixed, age dependent) | 6 | See table S.11 | Departament d'Educació, GenCat[15] |
| Company size in 5 sectors Sanitary, Agriculture, Industry, Services, Construction | 10 | Gamma function $\mu, \sigma$: (0.1, 70), (1., 1.), (0.35, 36), (0.2, 50), (0.1, 70) | Directorio Central de Empresas[14] |
| Fraction of contacts in subgroups, global (Havel-Hakimi) | 1 | 80%, 20% | Assumed |
| Size of subgroups in classrooms, companies | 2 | 10, 7 | Assumed |
| Temporary agricultural workers, number, size of companies | 3 | 4143, 20 (Fruits), 5 (Vineyards) | Directorio Central de Empresas[14] and IDESCAT[13] |
| Social contacts (Poisson distribution, age dependent) | 16 | See table S.11 | Synthetic contact matrices[33] |
| Incidental summer contacts | 34 | See table S.10 | IDESCAT[13], PADRIS[8] |
| Number of 8h slots spent in leisure during (normality, strong confinement) | 4 | General population: (8,2); Nursing facilities: (4,0) | Assumed |
| Reduction effective duration of contacts in nursing facilities: 1/(number of residents)$^N$ | 1 | N = 0.4 | ref.[29] and 1$^{\text{st}}$ wave data |

**Table S.8.** Parameters of the model (part II).

| Category | Number | Parameters | Source |
|---|---|---|---|
| Use of public transportation | | | |
| Fraction of workers (Work) | 1 | 30% | Autoritat del Transport Metropolità (ATM) [39] |
| Fraction of pupils (School) | 1 | 20% | ATM[39] |
| Fraction of population (social activities) | 1 | 10% | ATM[39] |
| Average number of contacts per round trip (work/school) (fixed number) | 1 | 1.0 | Assumed |
| Average number of contacts per round trip (leisure activities) (fixed number) | 1 | 1.1 | Assumed |
| Confinement | | | |
| Date of 1st day of simulation | 1 | 25 days before March 16, 2020 starting date of first confinement | Diari Official de la Generalitat de Catalunya [40] |
| Effect of mask | 7 | See Table S.12 | Medical data[41],[42] |
| Calibration of mobile phones data | 10 | (0.6,0.8,0.8,0.6,0.95) and dates | Figure S.2 |
| Overall normalisation of leisure mobility | 1 | 0.97 | 1st fitted parameter |
| Vaccination | | | |
| Reduction of contagion probability: 1st dose mRNA-based | 1 | 47% | Medical data [22] |
| Reduction of contagion probability: 2nd dose mRNA-based | 1 | 92% | Medical data [22] |
| Reduction of contagion probability: 1st dose viral-based | 1 | 40% | Medical data [22] |
| Reduction of contagion probability: 2nd dose viral-based | 1 | 76% | Medical data [22] |
| Reduction of viral load emission per dose | 1 | 10% | Medical data [23] |
| Vaccination campaign | 35 | see Table S.13 | PADRIS data[8] |

**Table S.9.** Fraction of asymptomatic, moderately and severely infectious people depending on the age.

| Age (years) | Asymptomatic | Moderately infectious | Severely infectious |
|---|---|---|---|
| $< 15$ | 0.67 | 0.27 | 0.06 |
| 15 – 64 | 0.39 | 0.44 | 0.17 |
| $> 64$ | 0.05 | 0.24 | 0.71 |

**Table S.10.** Parameters used to model additional summer contacts.

| Parameters | Value |
|---|---|
| Average number of additional contacts | 1.0 – 3.0 |
| Number of periods covered | Summer (up to 3 intervals), Holidays (3 periods) |
| Number of regions described | Barcelona (3), (Lleida (1), Tarragona (1) Girona (1) |
| Factor of increase between 2021 and 2020 | 2.5 |

**Table S.11.** Number of children per classroom in each age group and average number of stable social contacts depending on the age.

| Classrooms' size | | Number of social contacts | |
|---|---|---|---|
| Age | Size | Age | Mean |
| 0 | 7 | $< 5$ | 3.21 |
| 1 | 12 | 5 – 10 | 3.88 |
| 2 | 16 | 10 – 15 | 5.27 |
| 3 – 5 | 22 | 15 – 20 | 5.97 |
| 6 – 11 | 25 | 20 – 25 | 5.26 |
| 12 – 18 | 28 | 25 – 30 | 4.33 |
| | | 30 – 35 | 4.35 |
| | | 35 – 40 | 5.51 |
| | | 40 – 45 | 6.48 |
| | | 45 – 50 | 4.94 |
| | | 50 – 55 | 5.26 |
| | | 55 – 60 | 5.26 |
| | | 60 – 65 | 5.06 |
| | | 65 – 70 | 4.05 |
| | | 70 – 75 | 4.89 |
| | | $> 75$ | 2.95 |

**Table S.12.** The effect of wearing a mask is simulated by introducing a factor $F_{\text{Mask}}$ that reduces the viral load transmission at work or school and in leisure activities. As of October 17, 2020, a relaxation $F_{\text{MaskWearing}}$ of the discipline in the use of masks by young adults is assumed.

| Use of mask | Viral load reduction |
|---|---|
| Encouraged (27/4/20) | $F_{\text{Mask}} = 0.70$ |
| Imposed (18/5/20) | $F_{\text{Mask}} = 0.35$ |
| Relaxed use (from 13 to 35 year-old) | $0.35 \times F_{\text{MaskWearing}} = 0.80$ |
| Starting date of relaxation | October 17, 2020 |
| Isolation measures of nursing facilities residents | 0.5 |
| More contagious work contacts nursing facilities workers | 3 |
| Improved effectiveness mask hospital workers (1/7/20) | 0.33 |

**Table S.13.** Parameterisation of the age category, vaccine type, starting date, delay between first and second dose, fraction of population vaccinated for the first and second dose, respectively.

| Age category | Vaccine | Start Date | $\Delta$t doses 1-2 | Dose 1 | Dose 2 | $\Delta$t doses 2-3 |
|---|---|---|---|---|---|---|
| Sanitary | BioNTech | Jan 3 | 28 days | 100% | 100% | 7 months |
| 0-11 years | BioNTech | – | – | – | – | – |
| 12-19 years | BioNTech | Jun 7 | 28 days | 70% | 55% | 7 months |
| 20-34 years | BioNTech | May 21 | 28 days | 80% | 60% | 7 months |
| 35-44 years | BioNTech | Apr 28 | 28 days | 85% | 65% | 7 months |
| 45-54 years | BioNTech | Apr 4 | 28 days | 90% | 75% | 7 months |
| 55-59 years | BioNTech | Mar 28 | 28 days | 90% | 80% | 7 months |
| 60-64 years | AstraZeneca | Mar 7 | 56 days | 92% | 80% | 6 months |
| 65-69 years | AstraZeneca | Mar 1 | 56 days | 92% | 85% | 6 months |
| 70-74 years | BioNTech | Feb 20 | 28 days | 92% | 85% | 7 months |
| 75- years | BioNTech | Jan 17 | 28 days | 95% | 95% | 7 months |
